# Kidney research (APOL1 and FSGS) violates new ethics of ‘race’: Medicontextualism debunks bad science undermining healthcare

**DOI:** 10.1101/2024.09.15.24313684

**Authors:** Cyril O. Burke, Joshua Ray Tanzer, Leanne M. Burke

## Abstract

**BACKGROUND:** Apolipoprotein L1 (APOL1) gene variants, associated with increased risk of focal segmental glomerulosclerosis (FSGS) and kidney failure, were found in US patients of West African heritage but absent in European and Taiwanese patients—Taiwan having one of the highest rates of kidney disease. Methodological limitations could affect these findings. A history of segregation in the southern US led researchers to propose biologic ‘race’ as the root of disparity in kidney disease. We hypothesized that persistent US segregation racializes health, education, employment, and access.

**METHODS:** First, for cross-cultural clarity, we defined terms to orient international colleagues to the semantics of socially constructed ‘race’ (in Europe) and racialization of health (in the US). Second, we removed ‘race correction’ from estimated glomerular filtration rate under more equal conditions of US veterans. Third, we used data re-analysis to suggest alternative mechanisms for links between the medicontexts of socially constructed ‘race’, apolipoprotein L1 gene variants, and racialized kidney disease.

**RESULTS:** There was no ‘racial’ disparity in kidney failure under more equal social conditions. Geographically localized “ancestry markers” may proxy for ‘race’. ‘Colorism’ may be associated with apolipoprotein L1 variants linked to medicontextual (e.g., pigmentary) genes that may be tracked with a polygenic risk score (PRS) for structural racism.

**CONCLUSIONS:** Under equal conditions, comparable outcomes should be the expected norm for all, regardless of socially constructed ‘race’, ethnicity, or nationality. Medicontextualism locates disparity between socially constructed categories in disparate social conditions. Ending silence on ‘race’ debunks bad science (BS) and suggests potential insights from a “structural-racism PRS”. International collaboration to discourage misuse of socially constructed ‘race’ in healthcare and medical research is an actionable Population Health initiative with potentially high impact but low effort and cost.

**Graphical Abstract:** 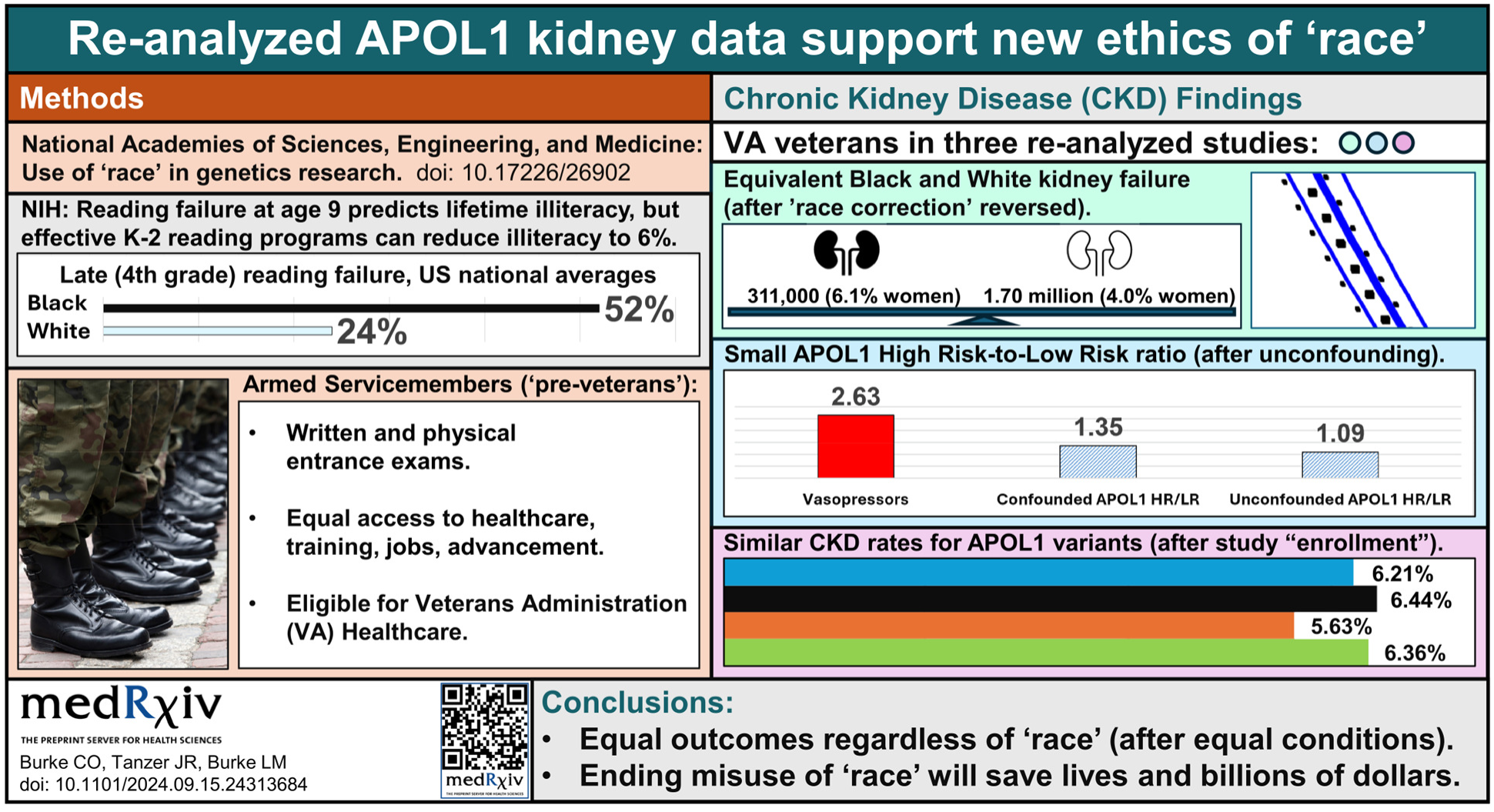

**Lead author’s preface:** Readers can judge our deference to reviewers in four rounds of review at one journal (posted in the 6 Comments to Version 9 of our preCKD article [1]). We submitted to journals in general medicine, internal medicine, nephrology, biomedical science, public health, diagnostic medicine, and ethics. “Peer” reviewers rejected the discussion of ‘race’ required by medicontextualism. We incorporated medicontextual explanations of ‘race’ to inform those unfamiliar with the roots of US ‘racial’ disparities. Several editors offered publication only <u>if</u> we adopted their view on ‘race’ [2], **Fig 0**.

Fig 0.
Medicontextualism for ‘racial’ disparities research suggested blind spots in academic “peer” review of ‘race’.

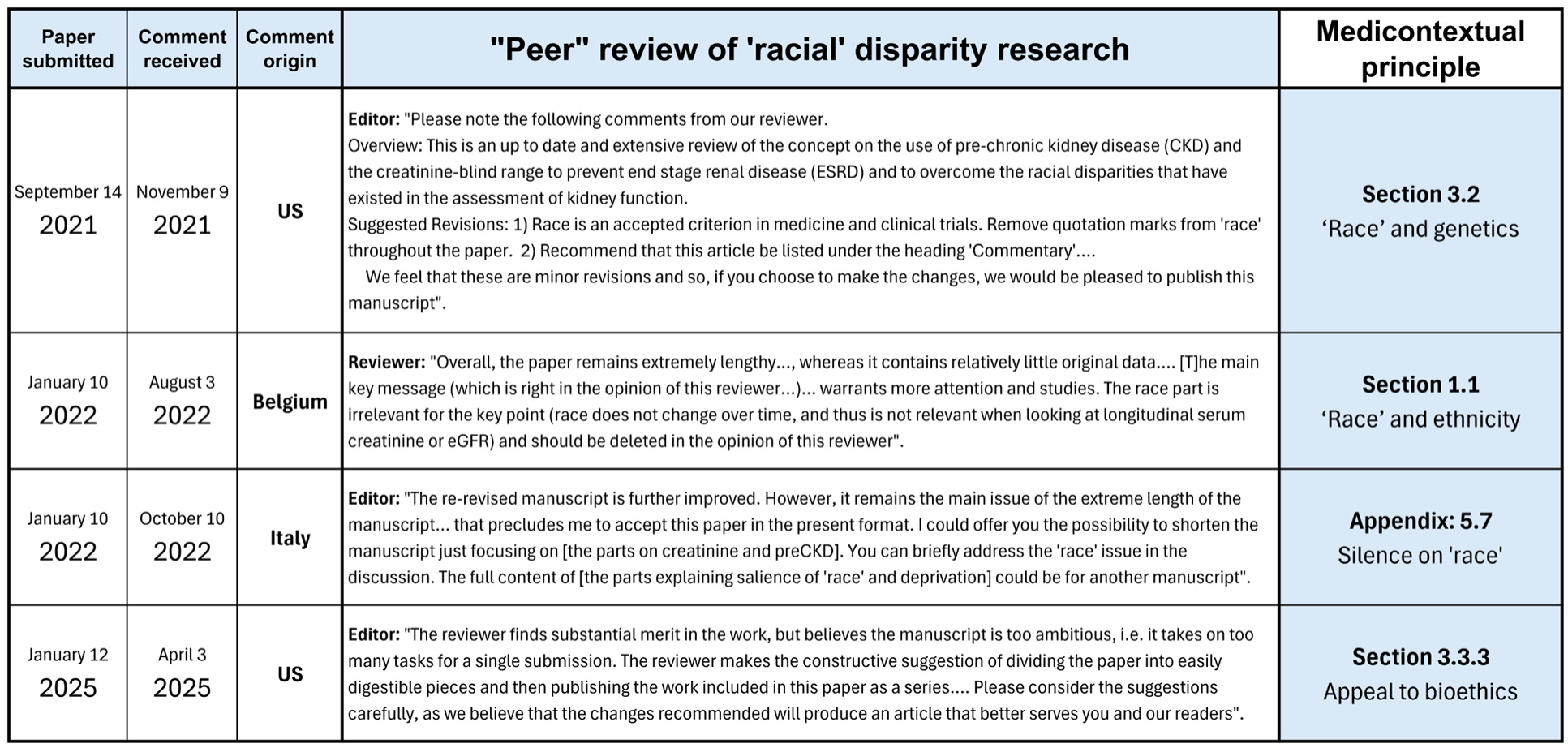

Our 3,000-word research manuscript progressively lengthened to present an alternative view of ‘race’. A US editor kindly proposed a $20,000 (in publishing fees) series on the ear, feet, tail, trunk, tusks, but frugality favored a single article for interested readers, including all the parts to reconstruct the medicontextual elephant in the room.

Pinker wrote, “…academia seems innocent of a basic phenomenon in social psychology: Closed circles of like-minded people can develop subcultures with entrenched norms and blind spots” [3]. Lentin described the origins of European silence on ‘race’ (see **Appendix: 5.7**).

## 1. INTRODUCTION

Medical research has long incorporated ‘race’ into clinical practice. The long history of misusing ‘race’ as an input variable in severity equations is joined by modern genomic associations, and more, couched in population differences by ‘race’. Missing from the discussion are the medicontexts that link causal factors to physical characteristics of individual ancestry. Persistent ‘racial’ segregation racializes US health, education, employment, and access. Not adequately accounting for the medicontext promotes what we call bad science (BS). We use the example of ‘racial’ disparity in US chronic kidney disease to discuss this complex topic, framed in what we call ‘medicontextualism’—the broad health and social environments (medical contexts or ‘medicontexts’) constructed to target physical characteristics of individual ancestry.

Unchallenged for ‘racial’ inconsistency and selectively including or overlooking its influence, medical research promotes ‘racial’ illusions that advance the myth of ‘race’ as biology. **Linda M. Hunt, Professor of Anthropology at Michigan State University** et al noted:

> …current health literature encourages clinicians to see their patients through a racial lens…. [T]he systematic inequalities that have been observed in the clinical care that racialized patients receive… likely reflect more than structural inequalities alone. They may also be attributable to this systematic authorization of race-based care. The arbitrary and contradictory ways the clinicians in this study understand and apply the concept of race raise concern that not only does using race to inform diagnostic and treatment choices lack scientific rigor, it may also put patients at risk of receiving nonstandard care. [4]

Variants of the *APOL1* gene were associated with kidney disease in the US and West African countries but “extremely rare” in Europe [5] and absent (despite some of the highest rates of kidney disease) in Taiwan [6]. We discuss how methodological limitations produce such inconsistent findings and ways ‘race’ shapes medical practice in the US and internationally. This **Introduction** provides examples to orient international readers to the illusions of socially constructed ‘race’ and ‘ethnicity’ in Europe and the US. Our **Results** are reanalyzed published data to illuminate observed differences that inspire bad science (BS) in racialized countries. Finally, after 100 years of failed efforts to end misuse of ‘race’, our deliberately comprehensive **Discussion** provides research and overview in a single article to reveal the elephant in the room.

### 1.1 ‘Race’ and ethnicity

Over time, the socially constructed meaning of ‘race’ changes. Hidden assumptions and substitution of “ethnicity” and other euphemisms may limit understanding by international colleagues. In *The meaning of racial or ethnic origin in EU law*, the European Commission noted:

> In EU law, the Racial Equality Directive (RED) prohibits discrimination on the grounds of racial or ethnic origin without defining them…. In Europe, most groups targeted by racism categorically reject race as a label. Still, racialisation as ‘a process that ascribes physical and cultural differences to individuals and groups’ is thriving….
>
> Without real physical differences, people can be racialised on the basis of their perceived religiosity: the dress they wear or the way they cover their hair. Theories of Christianity’s superiority over the Jewish and Muslim faiths can be traced back over centuries of European civilisation, and they laid the ground for anti-Semitism and Islamophobia. Poverty, too, can be portrayed as having its own culture, language and value system, thus poverty can also be racialised – particularly if a visible proportion of poor people belong to racial minorities….
>
> The traumatic experiences of the Holocaust and the urge to respond to apartheid provoked the scientification of race in legal terminology, which trapped racial in the confines of biological racism – the essence of the ideologies it originally set out to combat. Scientification disassociated racial from nation(al minority) and consequently severed cultural, religious and linguistic from it. Reducing the meaning of racial to ‘biological’ inevitably paved the way to essentialisation, while insulating it from the influence of social sciences….
>
> The invention of a supercategory with the intention of ensuring terminological unity and dispelling scientific misconceptions of race and racial origin at the international level had an unintended side effect in Europe. Here, the prism of nationalism resists the smooth transfer of terminological reform that followed WWII. While in **France**, ethnic origin is perceived as equivalent to race, in the **UK** it is separately defined in case law. In the rest of the EU, correlations between ethnic and racial occupy a central place in academic discussions and judicial interpretation. [7]

Uncertainties in socially constructed ‘race’ include **(1)** lack of “consensus definition of race or ethnicity”, **(2)** inconsistency between subject and observer identification for other than Black or White ‘race’, and **(3)** “fluidity” of self-identification [8]. Because co-located subject and observer are confident in the local meaning of Black and White ‘race’, their agreement reflects ‘shared illusion’ rather than scientific “…validity” [9,10,11,12,13,14]. And ‘race’ perceived by perpetrators of racist acts may correlate better with ‘racial’ disparities than self-reported ‘race’ [15].

In a 2023 report on *Using Population Descriptors in Genetics and Genomics Research*, the National Academies of Sciences, Engineering, and Medicine (NASEM) stated:

> Race is neither useful nor scientifically valid as a measure of the structure of human genetic variation….
>
> Over the past several decades, a number of professional societies have developed statements or guidelines on race, ethnicity, human diversity, and multicultural practices…. In every case, the guidelines have been aspirational and intended to encourage their professional colleagues to become educated and informed….
>
> Despite the many recommendations, guidelines, and strategies… there has been relatively little change in how any entities within the genetics and genomics research ecosystem use these descriptors or require them to be used. [16]

Medicontextualism exposes bad science (BS) derived from inconsistent, location-dependent definitions of ‘race’, lacking international reproducibility and generalizability.

### 1.2 Racialized US healthcare

Kidney disease is a worldwide public health problem with significant differences between ‘racialized’ groups. Although US healthcare spent more per capita than any other nation [17], it ranked last among high-income countries [18], and 48th of 50 for kidney-related mortality [19], **Fig 1**.

**Fig 1.**
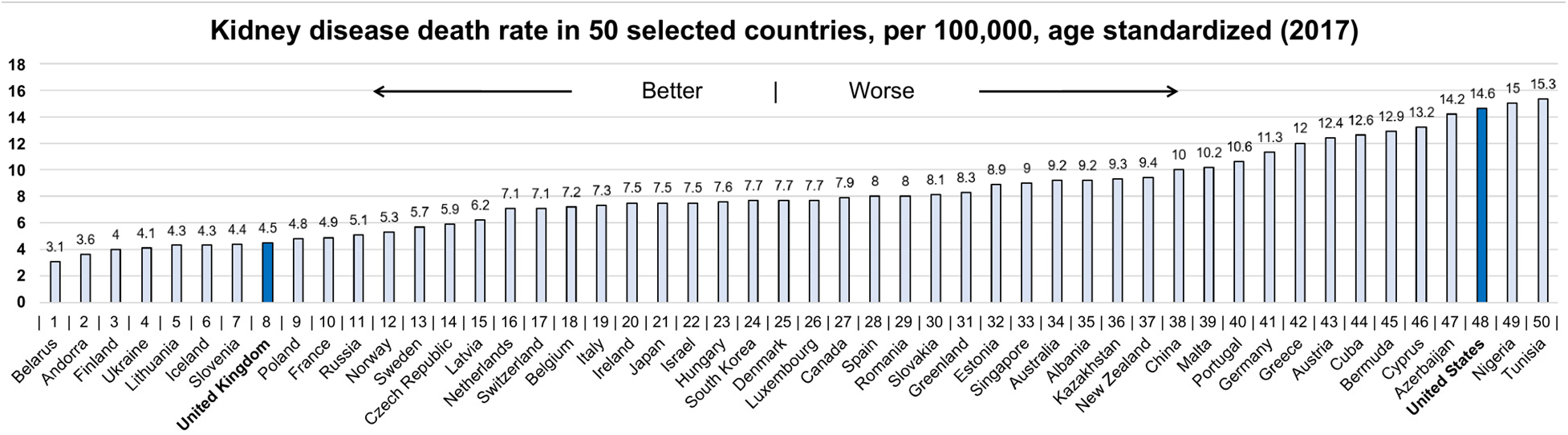
Age-standardized kidney disease death rate per 100,000 in 50 selected countries (2017).

Federal spending for chronic kidney disease (CKD) and kidney failure (KF) [20] doubled in a decade, reaching $122 billion [21], or 1.8% of total federal spending, **Fig 2**.

**Fig 2.**
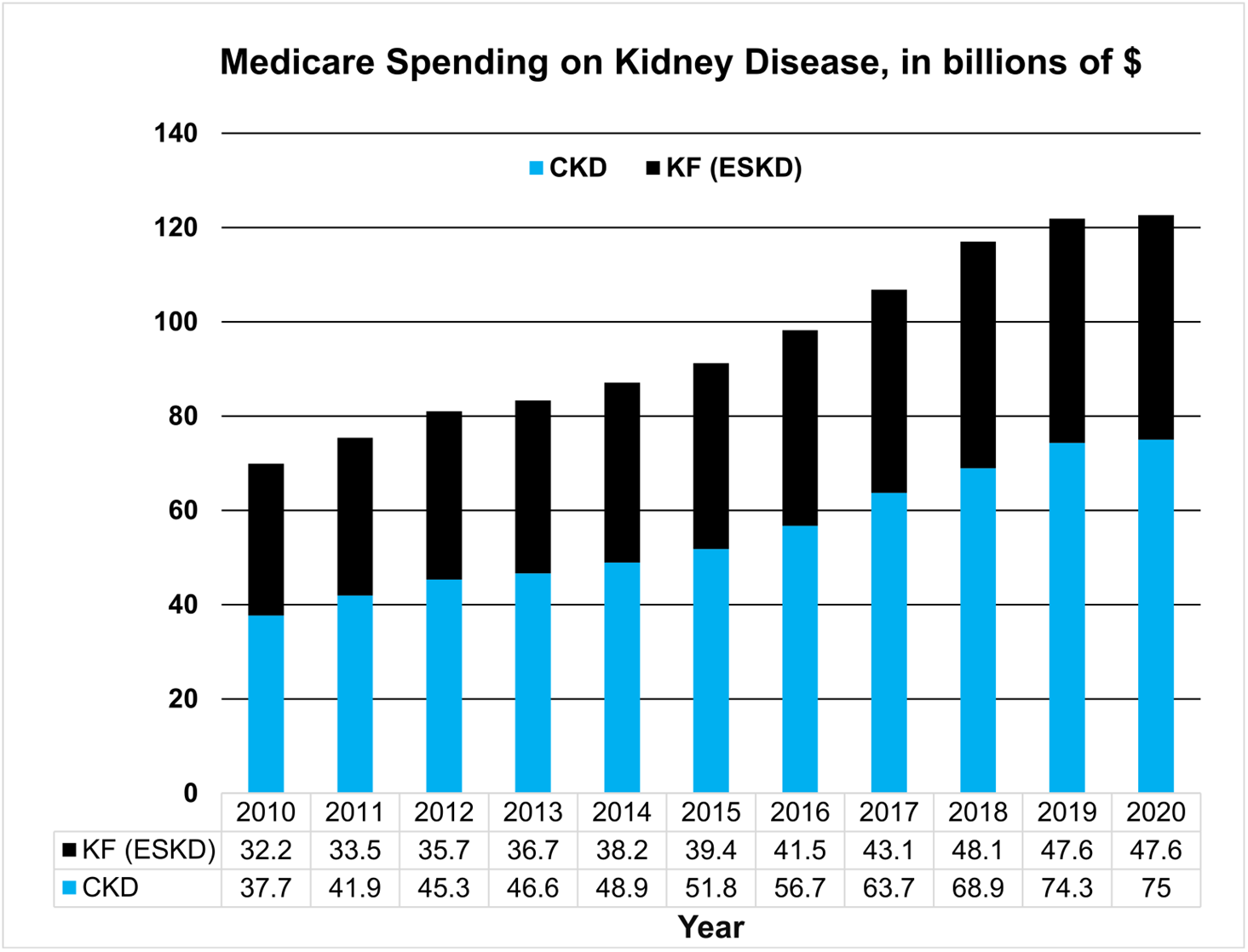
Medicare spending on chronic kidney disease (CKD) and kidney failure (KF).

#### 1.2.1 ‘Racial’ profiling, hypertension, kidney disease

Unless the underlying, medicontextual mechanisms of inequality are directly measured, recruiting ‘racially diverse’ subjects into medical research will find ‘race’ in disparate treatment from lingering ‘racial’ segregation and structural racism [22,23].

In the US, CKD and KF represent the eighth leading cause of death, with lifetime risk varying by socially constructed ‘race’ [24,25], **Fig 3**. US CKD is “…one of the starkest examples of racial/ethnic disparities in health” [26]. Black Americans are 12% of the population but 35% of patients on dialysis. In contrast, foreign-born Black and White persons “…had a lower likelihood of CKD than those US-born”, with foreign-born Black adults at 51% lower risk than Black Americans [27]. Ending ‘racial’ disparity in US kidney disease for Black Americans <u>alone</u> could redirect 20% of federal funds—$25 billion annually—from late-CKD to prevention [28]. Excess CKD also affects Hispanic, Asian, Native American people, and others.

**Fig 3.**
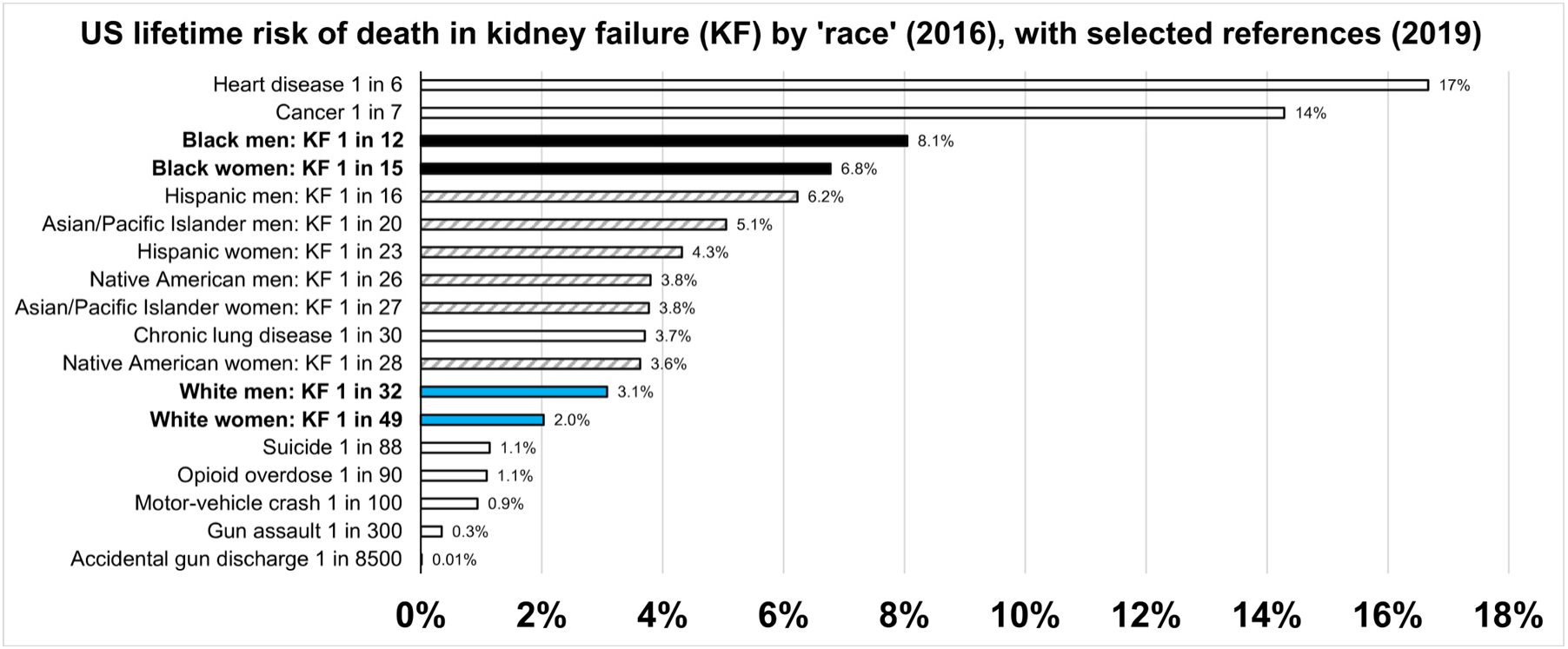
Lifetime risk of death in kidney failure (KF), with selected references. Solid and striped bars show KF death rates by US ‘race’ and ethnicity. Clear bars show death rates for other causes (combined sexes).

US kidney disease mortality (per 100,000 per year) varies by city, ranging from 2.0 in San Diego to 18 in Indianapolis for White Americans; and from 7.9 in New York City to 45 in Charlotte, North Carolina for Black Americans [29]. The <u>ratio</u> of Black-to-White kidney mortality shows a different geographic pattern, **Fig 4**. Kidney dialysis outcomes are worse “…in regions with lower income, higher unemployment, … lower college education, and… greater proportions of Black and Hispanic residents…. concentrated in the southern and western United States” [30]. Life expectancy also varies with location and ‘race’ [31,32,33].

**Fig 4.**
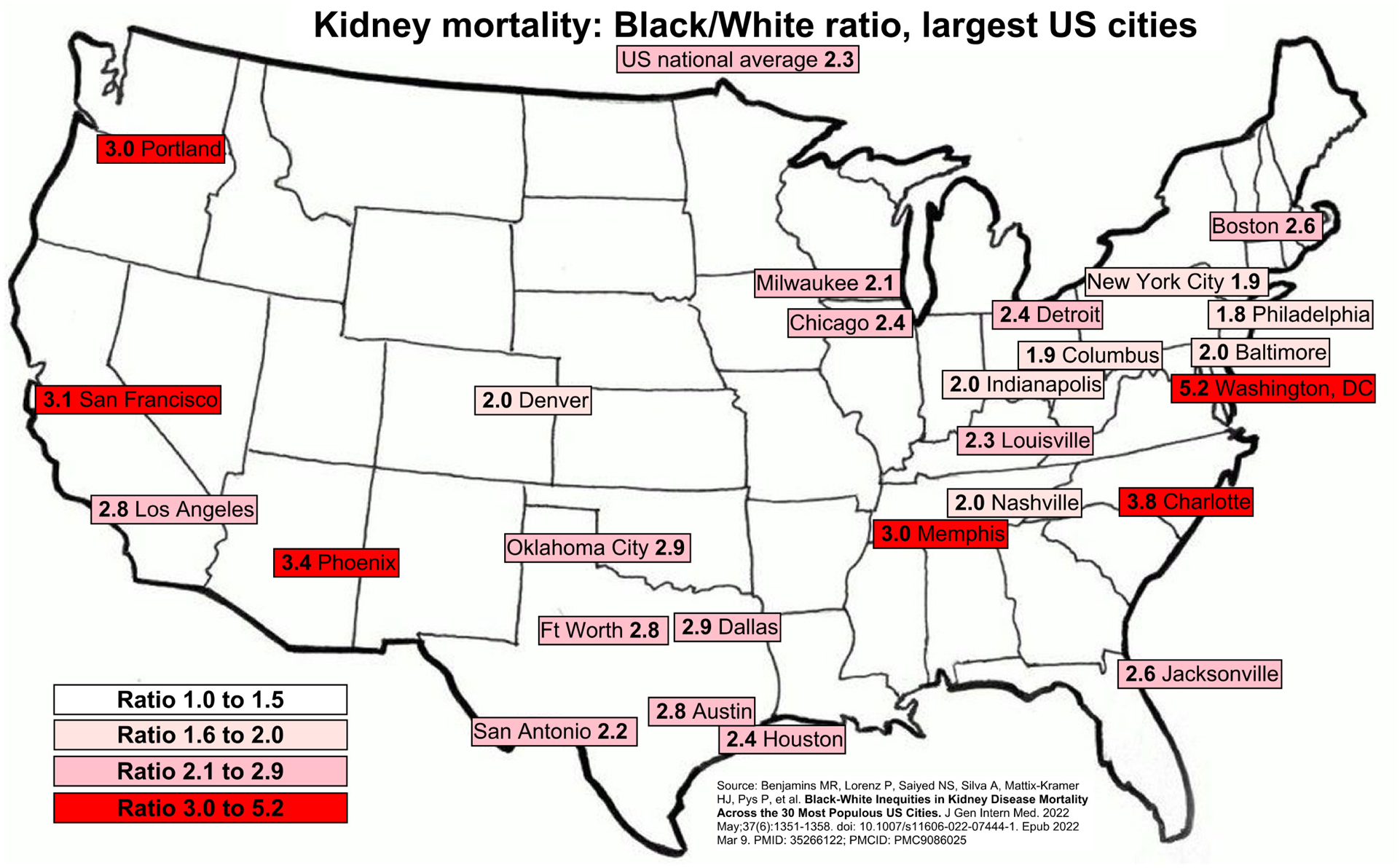
Ratio of Black to White mortality from kidney disease in largest US cities (2014-2018).

There are numerous “…racially segregated US hospital markets” [34]. “Low-income minorities with bad health had 68% less odds of being insured than high-income Whites with good health” [35]. At some academic centers, Black Americans were more likely to be deemed “teaching cases” for training the medical students, residents, and fellows. In 2003, the National Academy of Medicine reported on *Unequal Treatment: Confronting Racial and Ethnic Disparities in Health Care* [36], but 20 years later, “…There hasn’t been a lot of progress…. We are still largely seeing what some would call medical apartheid” [37].

Data collected to monitor ‘racial’ disparities can worsen them by misuse in clinical algorithms [38] and clinical decision-making tools [39,40]. **Hunt** and Kreiner noted the consequences for primary care: “…that the idea of ‘Personalized Medicine’ can so ironically be converted into carte blanche for practicing racialized medicine” [41].

Medical studies often omit medicontextual information relevant to ‘racial’ disparities. For example, dehydration is prevalent among Black Americans [42,43], causes more acute kidney injury (AKI) in low-income areas [44], and increases CKD risk [45,46]. US treatment guidelines for hypertension in Black patients favor thiazide diuretics [47,48] that can worsen dehydration. A US study recommended against diagnosing KF while on thiazides, noted thiazide-to-loop-diuretic usage ratios of 1.06 for White and 1.66 for Black subjects, but omitted KF by ‘race’ [49]. A US study recommended systolic blood pressure control to less than 120 mm Hg (to reduce cardiovascular mortality), reported no benefit in Black subjects [50], but omitted their 100% increase in AKI (until years later [51]). A US study recommended a thiazide for poorly controlled hypertension in advanced CKD, reported that chlorthalidone tripled the number with AKI and increased AKI episodes per subject by 50%, but omitted AKI by ‘race’ and that it used a ‘race-corrected’ eGFR that underestimates CKD and its severity in Black people [52].

#### 1.2.2 ‘Race correction’

One way ‘race’ is codified in medical practice is ‘race correction’. The first ‘race correction’ for estimating glomerular filtration rate (GFR) led to more estimated GFR (eGFR) ‘corrections’—for ethnicity in the US [53] and nationalities in Asia [54,55,56,57]. ‘Race corrected’ eGFR results led to questioning [58,59], and the “…majority of studies found that removal of race adjustment improved bias, accuracy, and precision of eGFR equations for Black adults” [60]. Some condemned the “…inappropriate use of race in nephrology” and distinguished mistreating individuals from monitoring the effects of racism on a population [61]. Others hailed “race-free” equations as a “…successful compromise” that removed ‘race’ only from the equations [62]. Reports that “race-free” eGFR equations “…underestimated measured GFR in Black participants… and overestimated measured GFR in non-Black participants” [63] merely shifted misuse of ‘race’ to pre-test “population weighting” [64,65], potentially delaying care for Black patients [66] in a way much harder to detect and stop.

#### 1.2.3 Science and outcomes

The problem with ‘race’ in medical research relates largely to methodology. Many view scientific research as objective, providing factual answers to questions posed. However, the relevance and rigor in testing an hypothesis always have limitations, especially for something nuanced like ‘race’. The old adage, “garbage in, garbage out”, captures a fundamental problem with bad science (BS).

Countering the narrative of objectivity, Thomas S. Kuhn, a preeminent scholar on the philosophy of science, presented ‘science’ as inherently social [67]. Yet, medical research examining socially constructed ‘race’ as a proxy for social determinants of health (SDOH) is often demoted to social ‘Commentary’ [2]. Articles proclaiming the objectivity of quantitative analysis [68,69] miss how confirmation bias influences what numbers we collect, how we understand that data, our confidence in what we are doing now, and which articles get published. After 100 years of academic articles that ‘race’ is a myth, ending misuse of ‘race’ is an overdue Kuhnian paradigm shift [67,70,71].

“With malice toward none, with charity for all” [72], we re-examined ‘race’ in kidney disease. Removing ‘race correction’ from GFR estimates of US Black and White veterans showed comparable results. Re-analyzing two studies in US Black veterans of gene variants associated with kidney risk—both published before NASEM guidance [16]—gave insights into the medicontextual influence of socially constructed ‘race’.

## 2. RESULTS (three secondary data re-analyses)

### 2.1 Removing ‘race correction’

To show the practical implications of the ‘race correction’, we first reanalyzed data to remove ‘race correction’ from GFR estimates. We considered an impressive US study of KF versus eGFR in 1.70 million White veterans (4% female) and 311,000 Black veterans (6.1% female) [73] to investigate the medicontextual public health implications and demonstrate that bad science (BS) comparisons between arbitrary groups would be less after comparable conditions of selection and opportunity.

#### 2.1.1 Equal outcomes despite ‘race’

The primary study, *White/black racial differences in risk of end-stage renal disease and death* [73], reported baseline US veteran characteristics and renal comorbidities (all significant to p<0.001), **Fig 5**, with Black veterans more likely to be younger, female, hypertensive, diabetic, in the highest and lowest eGFR ranges, and living in lower-socioeconomic-status zip codes **(left)**, the latter also shown graphically **(right)**.

**Fig 5.**
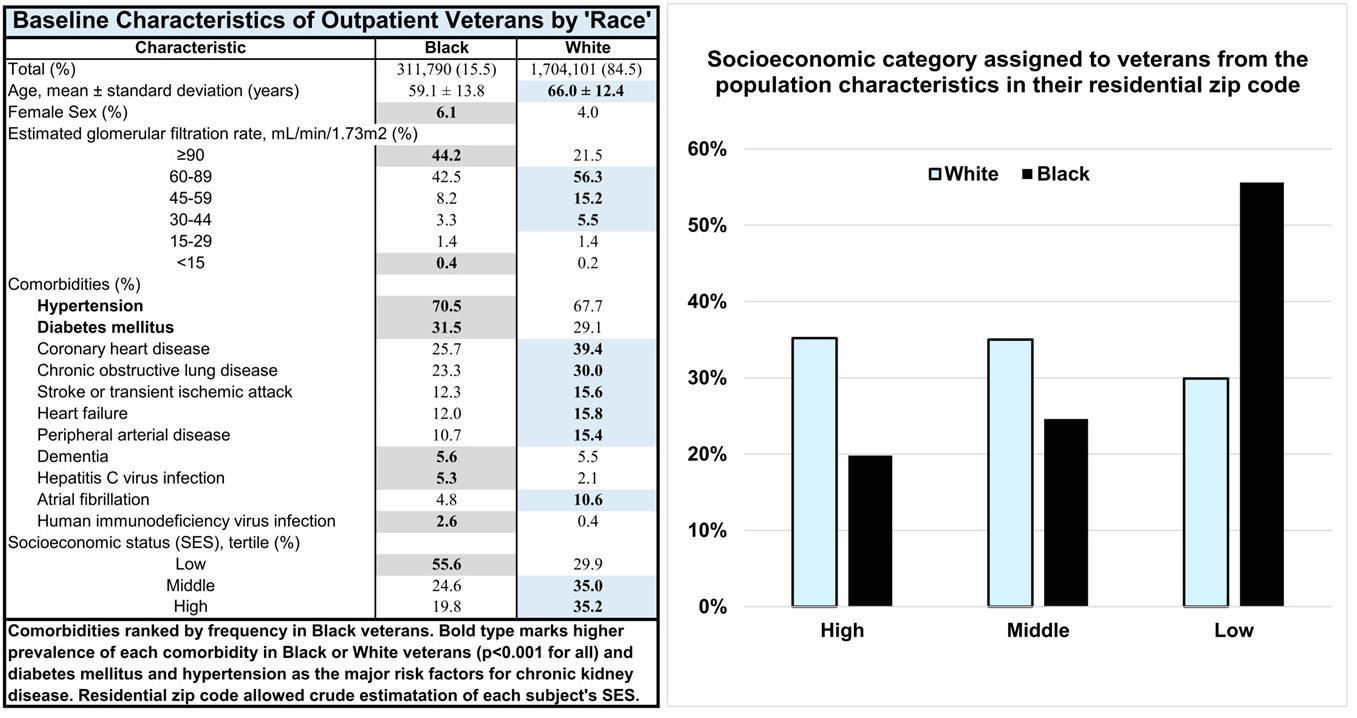
Comparing veteran characteristics by ‘race’ (left): veteran characteristics in the primary source by ‘race’, all cofactors with p<0.001, **(right):** veteran socioeconomic status, approximated in the primary source by zip code.

The Modification of Diet in Renal Disease (MDRD) eGFR ‘race correction’ for Black veterans can be removed by simply dividing by 1.21. Making this adjustment shifted rates for KF risk and eGFR prevalence to lower GFRs, **Fig 6**, also shown graphically in **Fig 7**. The X-axis ‘race corrections’ (horizontal arrows) sometimes had significant Y-axis effects (vertical arrows). The revised KF outcome curves (increasing to the left) overlapped for Black and White veterans (inset). The revised eGFR-prevalence curves (increasing to the right) showed higher prevalence of lower eGFRs in Black veterans. This shows how, under medicontextually similar conditions, the paradox of low prevalence of early CKD yet faster progression at late stages appeared to be a mathematical artifact of ‘race correction’.

**Fig 6.**
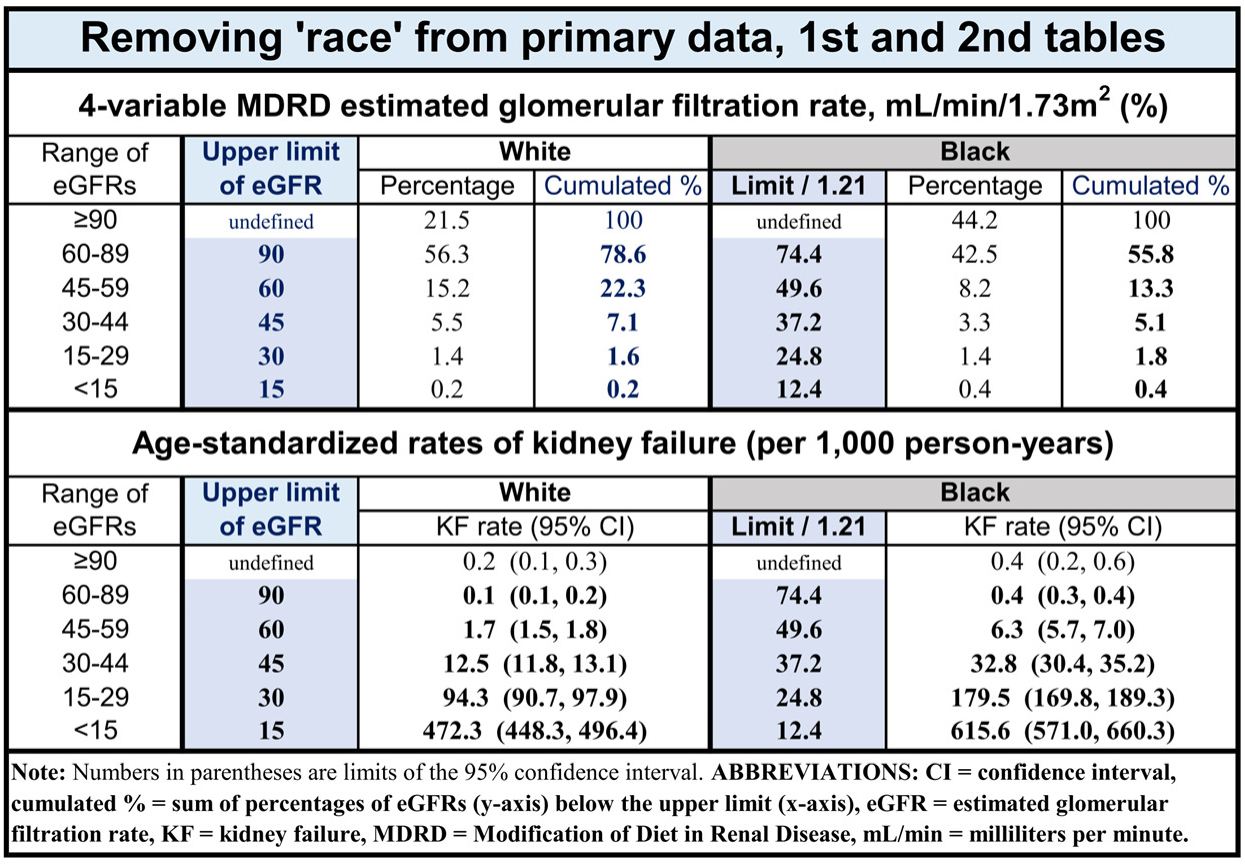
Removing ‘race correction’. Dividing the upper limit of eGFR range for Black patients by the 1.21 ‘race correction’ numerically removed ‘race correction’, shifting their results to lower numbers on the X-axis.

**Fig 7.**
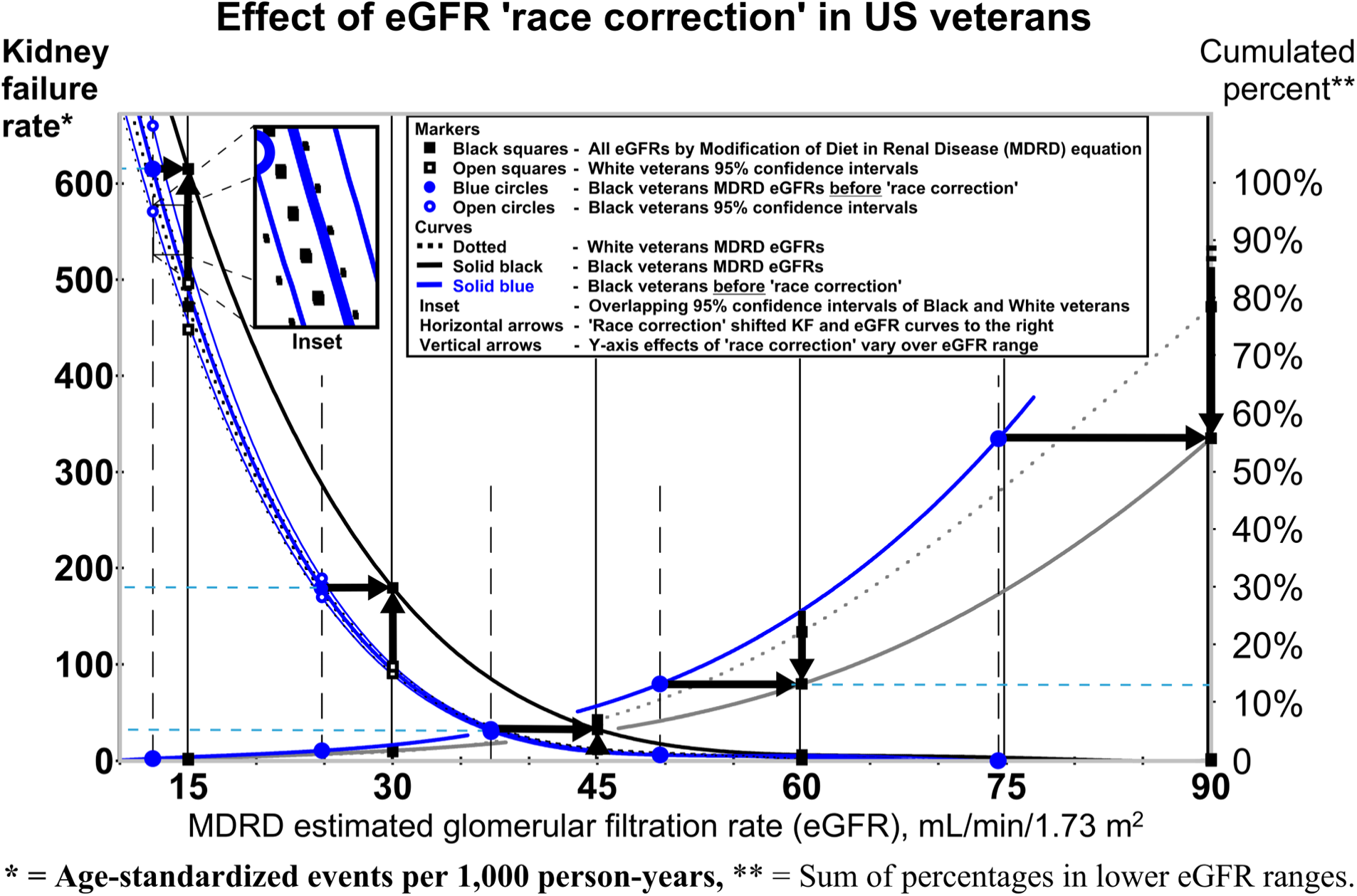
Removing ‘race correction’. Dividing by 1.21 shifted X-axis values to the left (blue curves). Horizontal and vertical arrows show X- and Y-axis shifts from ‘race correction’. **Curves increasing to the left:** kidney failure (KF) curves for Black and White veterans overlap within their 95% confidence intervals (inset). **Curves increasing to the right:** ‘race correction’ falsely lowered cumulated Y-axis eGFR percentages of Black veterans.

To further investigate, we intentionally removed the race correction to see how predicted levels of GFR matched actual data. When removing the ‘race correction’, White and Black patients had more similar associations between serum creatinine and GFR (*t* (10) = 0.91, *p* = 0.3860) than when ‘race correction’ was maintained (*t* (10) = 2.36, *p* = 0.0402). While the sample size is small and may not replicate in all settings, it shows how the ‘race correction’ is mathematically inducing differences that would not be identified if no ‘race correction’ were employed.

### 2.2 APOL1, AKI, and COVID

In an impressive primary study, *APOL1 Risk Variants, Acute Kidney Injury, and Death in Participants With African Ancestry Hospitalized With COVID-19 From the Million Veteran Program*, Hung et al examined the effects of apolipoprotein L1 (APOL1) risk variant (RV) alleles among 990 participants in the US Department of Veterans Affairs Million Veteran Program with “African ancestry” and hospitalized with coronavirus disease 2019 (COVID-19) [74]. It was suggested that *APOL1* gene variants protective against sleeping sickness in Sub-Saharan West Africa, and therefore more common in that area, also confer risk for KF. While we do not contest the prevalence of these variants in the African diaspora, it is less clear whether these variants are the mechanism for differences in medicontextual KF rates between racialized groups.

**Fig 8** shows AKI rates with and without various cofactors, ordered by odds ratios (ORs). The study found relative risk (RR) of AKI was 1.35 times higher for *APOL1* “high-risk” (HR) genotype (*APOL1* HR = 2 RV copies) than “low-risk” (LR) genotype (*APOL1* LR = 0 <u>or</u> 1 RV copy). However, RR of AKI was 2.4 for mechanical ventilation and 2.63 for vasopressor use, both more common in the *APOL1* HR group.

**Fig 8.**
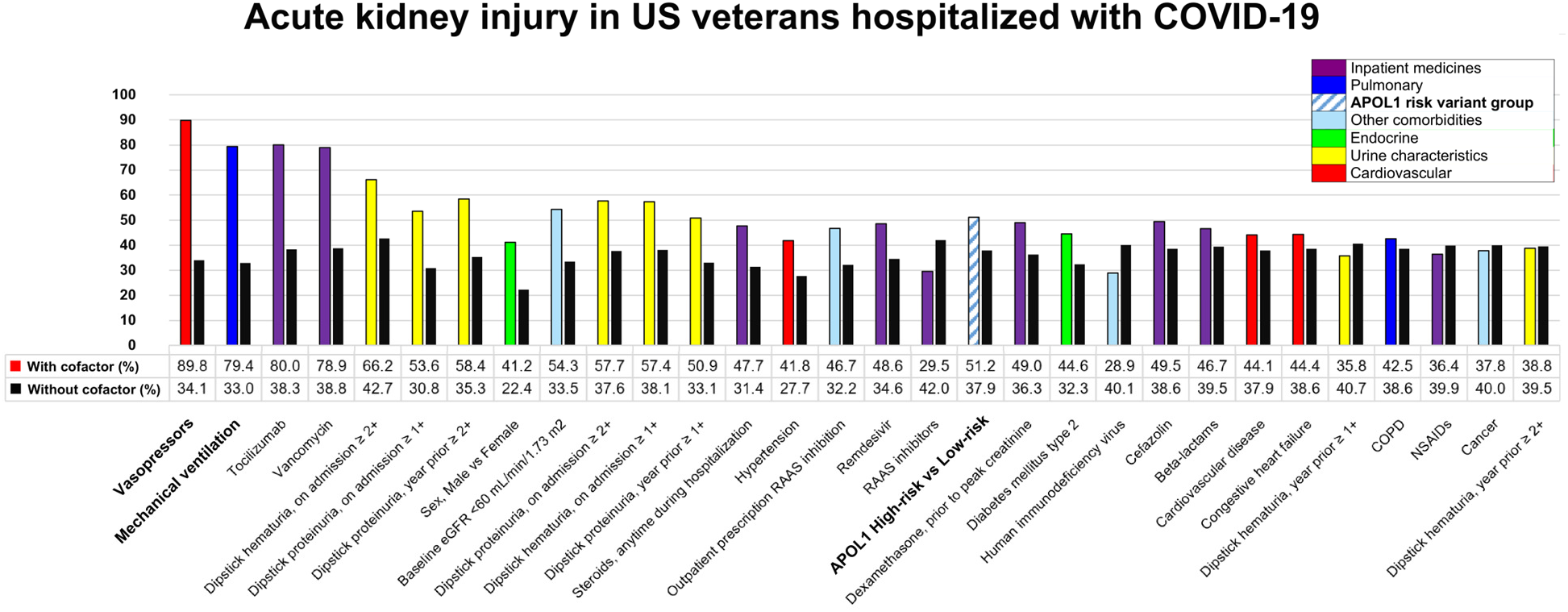
Acute kidney injury rates for various cofactors in veterans hospitalized with COVID-19. Possible direct or indirect AKI cofactors ordered by odds ratios (ORs), the greatest risk correlating with indicators of poor health.

Mechanical ventilation and vasopressors showed the largest RRs, consistent with findings that AKI in US veterans with COVID-19 varied with severity of illness [75]. Verbeek et al recommended correcting bias in observed RR using the “reference confounder” with the largest effect [76]. Unconfounding the *APOL1* RR of AKI with vasopressors alone reduced the RR to 1.09, **Fig 9**.

**Fig 9.**
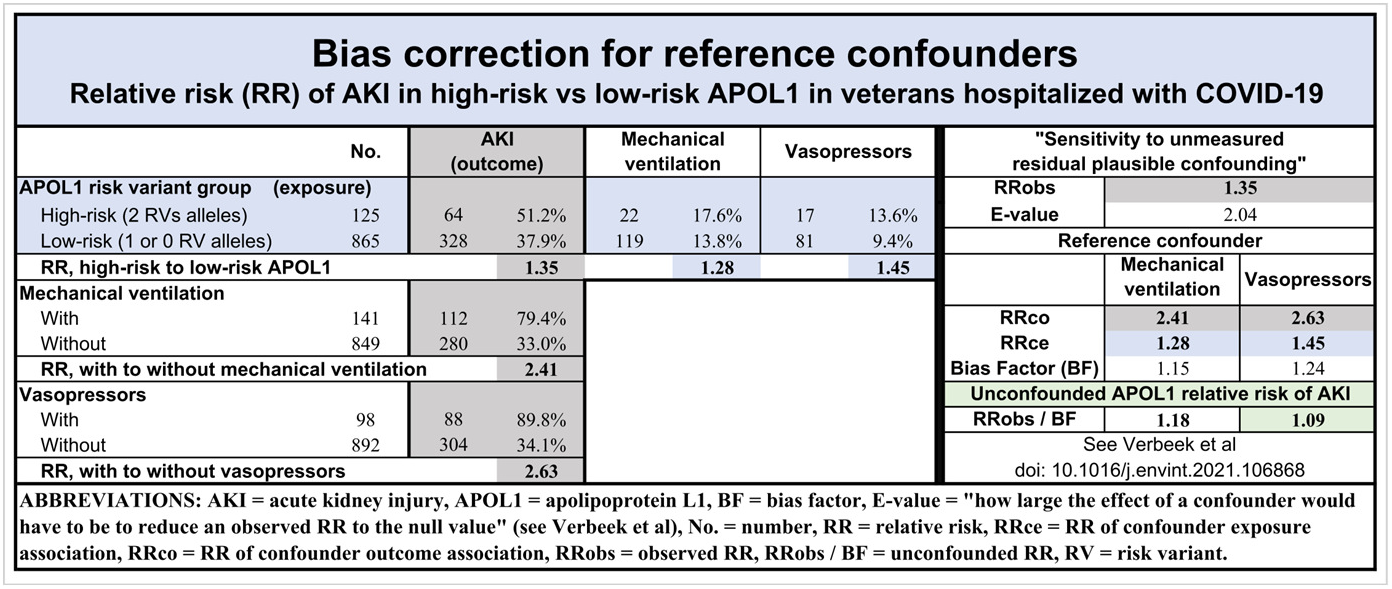
Bias correction for reference confounders. Correcting effects of vasopressors and mechanical ventilation on AKI outcomes reveals unconfounded relative risk (RR) of *APOL1* high-risk versus low-risk genotypes. Correlation between mechanical ventilation and vasopressors was not 100 percent, so unconfounded AKI risk would be even closer to unity on adjusting for both.

To ensure that these findings are not exclusive to the US, we compared available findings between the US and Brazil, results presented in **Fig 10**. This includes data from a similar study of COVID-19 and AKI in the Brazilian general population [77]. Although confidence intervals are much wider for the Brazilian sample due to a smaller sample size, the ORs between Brazil and the US are close for many cofactors.

**Fig 10.**
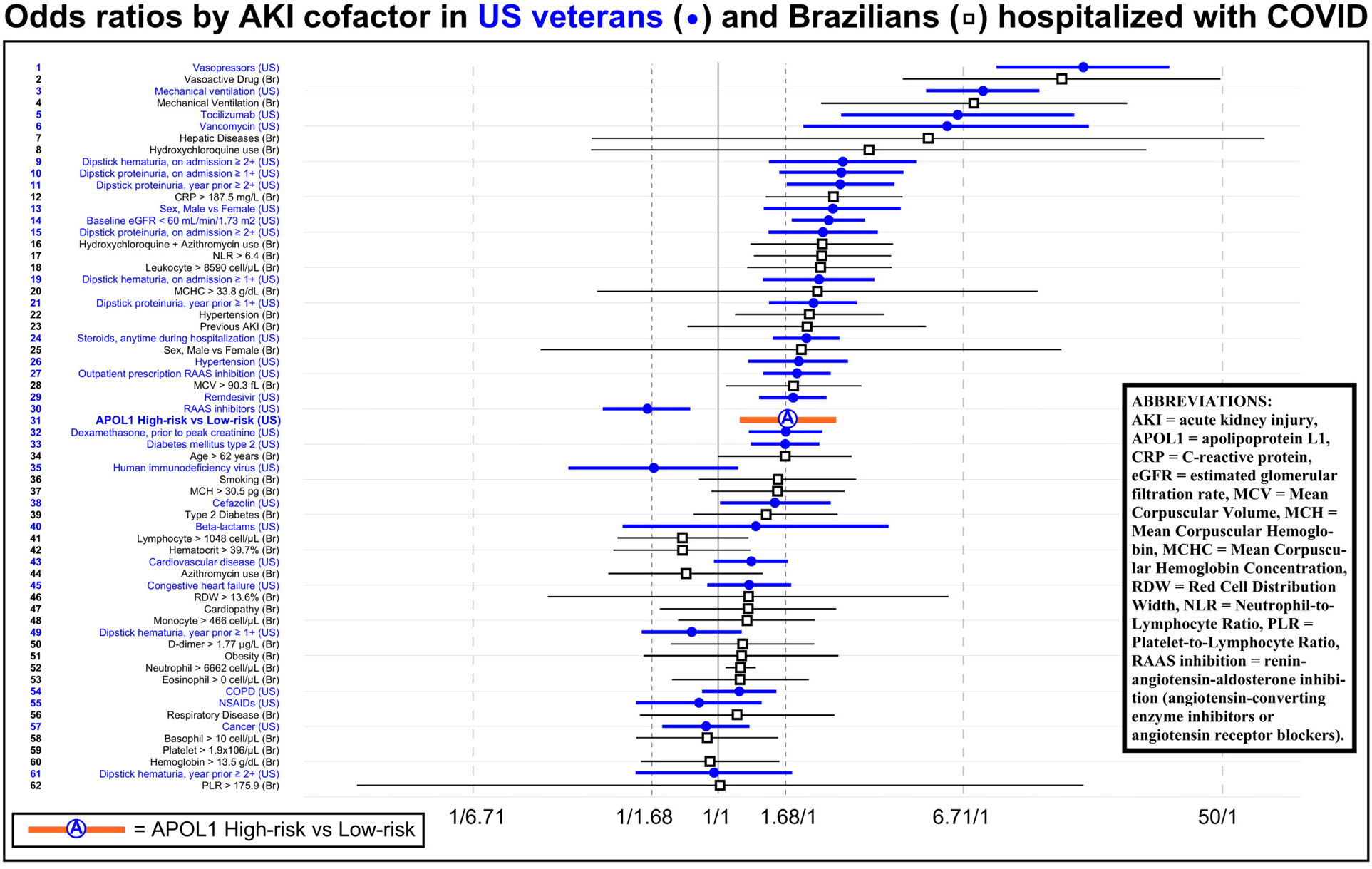
Acute kidney injury (AKI) odds ratios (ORs) for cofactors during hospitalization with COVID-19. Forest plot of AKI ORs (with 95% confidence intervals) suggests similarity of cofactors between US veterans and Brazilian general population despite differences in genetic origins and socially constructed ‘race’.

Many covariates showed statistically “small” effects (ORs below 1.68), some small enough to suggest random chance. Signs of critical illness showed “large” effects (ORs above 6.71). Urine characteristics had ORs primarily toward the upper range, highlighting their importance. The low OR for “dipstick hematuria, year prior” [rows 49 and 61] might reflect two sub-populations: women (some menstruating) and men (hematuria always abnormal). This shows how, in the greater medicontext of patient acuity, the RV allele of the *APOL1* gene does not characterize very large differences between racialized patient groups.

### 2.3 APOL1 and N264K

A second important primary study by Hung et al, *Genetic Inhibition of APOL1 Pore-Forming Function Prevents APOL1-Mediated Kidney Disease* [78], included 121,000 from the Million Veteran Program with “African ancestry” (13.8% female) and compared CKD in US patients with *APOL1* HR versus LR genotype and with and without N264K (a second-site *APOL1* variant). The study concluded, “…presence of a single copy of the *APOL1* p.N264K mutation mitigated the increased risk conferred by two *APOL1* HR variants”. About 20% of their data came from smaller studies in the general population, but our re-analysis used only 80% of their data, from US veterans.

**Fig 11** shows the percentage of subjects by kidney outcomes for four combinations of *APOL1* HR or LR and N264K alleles present (**+**) or absent (**-**) [78]. Black bars, representing *APOL1* high-risk genotype without N264K (*APOL1* HR, N264K**-**), show the highest rate for every kidney outcome. Blue bars represent the largest group, *APOL1* low-risk genotype without N264K (*APOL1* LR, N264K**-**). Note that presence of the N264K allele (N264K**+**) in just 3.8% of *APOL1* LR and 0.5% of *APOL1* HR patients means orange and green bars combined represent less than 5% of subjects, with small absolute numbers (population times rate) in their CKD and outcome groups.

**Fig 11.**
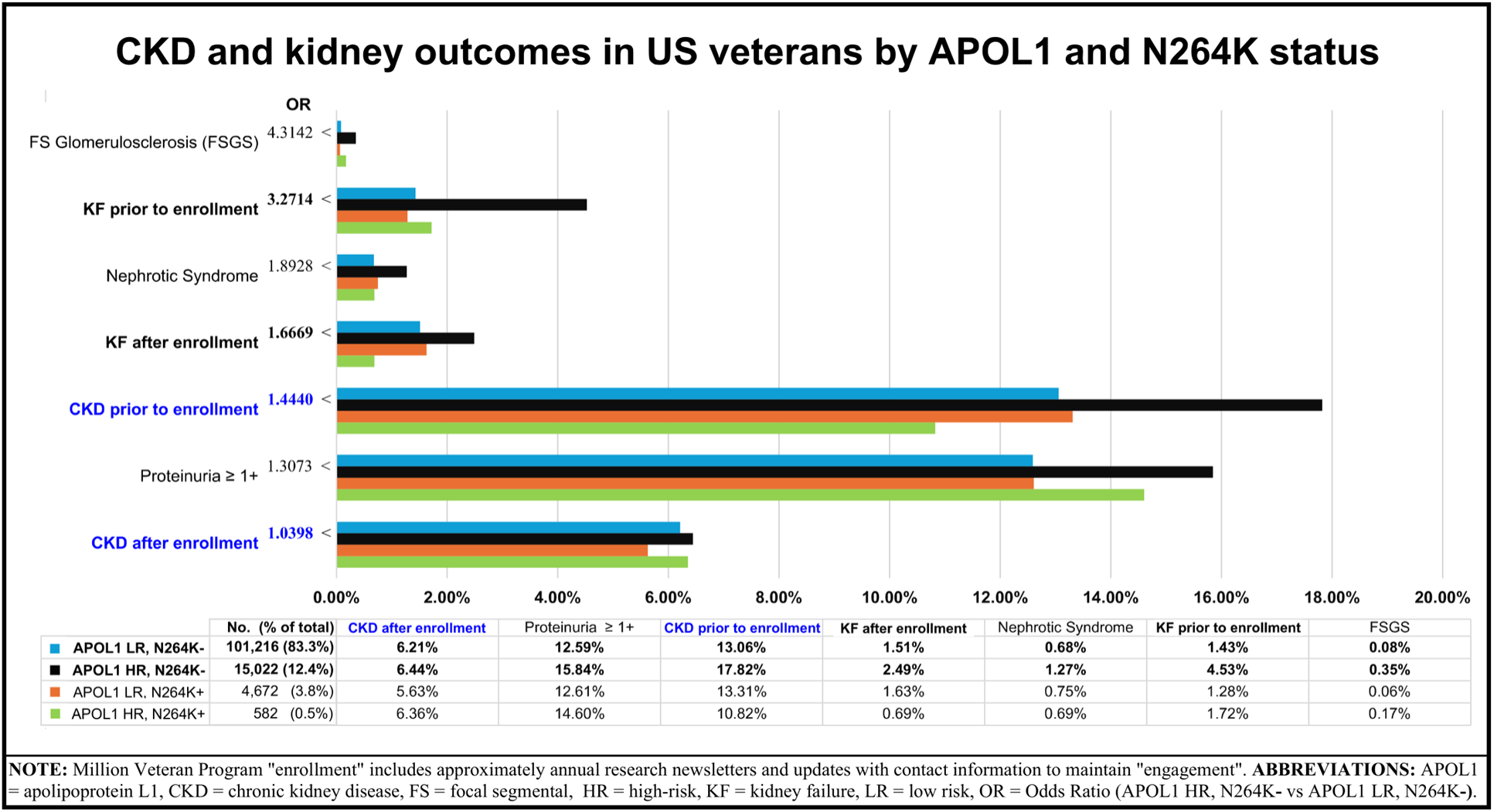
CKD and kidney outcomes in veterans by *APOL1* and N264K status. Ordered by odds ratio (OR) of kidney outcome between *APOL1* low-risk versus high-risk <u>without</u> N264K allele (N264K**-**)—less than 5% of subjects were positive for N264K (N264K**+**). Note the improvement in outcomes after study “enrollment”.

The chart is ordered by OR between the two largest groups <u>without</u> N264K (N264K**-**), representing 95% of Black veteran subjects and differing only by *APOL1* HR vs LR. Note for *APOL1* HR N264K**-** the association of lower rates of KF and CKD after than before Million Veteran Program “enrollment” [79] and that, after enrollment, rates of CKD were the same for all four groups. This is important because attributing KF and CKD to an inherent genetic problem can give the impression that there is limited ability to intervene. The similarity of CKD rates after enrollment suggests that all patients could benefit from treatment, even if there are differing acuity levels at baseline.

## 3. DISCUSSION

Stereotyped healthcare for ‘racialized’ patients promotes bad science (BS) through the most fundamental problem of research design: confounding. We cannot randomly assign personal heritage. It is impossible to separate personal experience from the way subjects look and the insults directed at their appearance. By ignoring persistent ‘racial’ segregation and ‘racial’ disparities that confound ‘racialized’ data, research perpetuates ‘race’ as biology, promotes interventions based on socially constructed ‘race’, and distracts from medicontextual public health conditions more relevant to population well-being.

To be clear, ‘race’ is a lingering social reality but a biological myth. Despite centuries of claims of biological difference, ‘race’ was absent under the high standards of the Cochrane Reviews. A study of 1,000 randomly selected Cochrane Reviews, published between 2000 and 2018, showed that only 14 (1.4%) had planned to include subgroup analysis by ‘race’ [80], and in those 14, there was only one racial’ finding of statistical significance.

Bioethical oversight of ‘race’ falls under a principle defined after decades of misuse of ‘race’ by the US Public Health Service [81,82], **Fig 12**. Yet, misuse of ‘race’ as a conscious or unconscious <u>input</u> for clinical care persists, even among those aware of its scientific invalidity [83]. Voluntary corrective guidelines “…have had little effect on how these concepts are deployed” [84]. Even the comprehensive, scholarly NASEM recommendations are voluntary [16].

**Fig 12.**
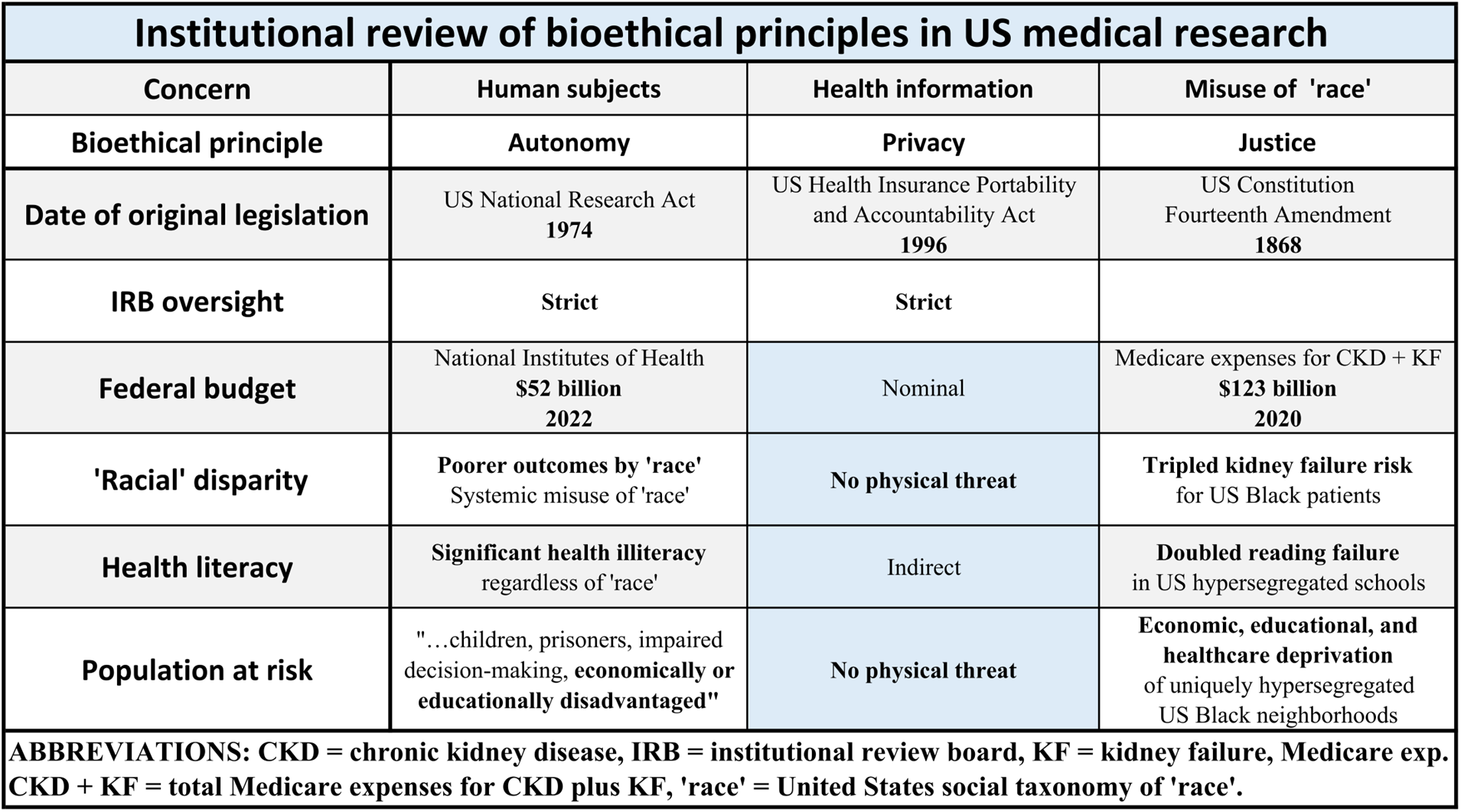
Bioethics: Autonomy, privacy, and justice in United States (US) healthcare. ‘Race’ collected to monitor progress in ‘racial’ disparities can be misused in clinical care. The principal of Justice could inspire oversight.

From any continent, studies under similar social conditions (e.g., even so-called “single race” studies) should be equally informative and relevant worldwide, regardless of the ‘race’ of the subjects (or lack of contrasting ‘races’). The lowest KF mortality rate (i.e, of the most privileged group in the ideal environment) should be the goal for all. Different outcomes reflect medicontextual social inequality.

To avoid perpetuating false notions of biological ‘race’ [85,86], we often label it “socially constructed” and always put ‘race’ or ‘racial’ in quotation marks [87,88].

### Ground zero for US kidney research

North Carolina (NC) has one of the highest rates of Black KF mortality and among the highest ratio of Black-to-White KF mortality, **Fig 4**.

In 1994, researchers in Chapel Hill, NC, proposed “collapsing glomerulopathy” (CG) as a new entity, with higher prevalence in Black patients, higher baseline CKD, and worse survival [89]. From only 16 cases between November 1988 and October 1990, they noted, “…this pattern of disease was rare in our region until recently and is now increasing in frequency”. They claimed, “No patients had known toxin exposure”.

In 2009, researchers in Winston-Salem, NC, hypothesized that kidney disease in “Self-reported African Americans born in North Carolina, South Carolina, Georgia, Virginia, or Tennessee”, previously attributed to hypertension, had genetic origin in the *MYH9* gene: “This gene exhibits impressive evidence for association with common, complex kidney diseases, with ORs as high as 3.4 for associated SNPs” [90].

Some researchers noted the possibility of non-genetic unknowns, but overall, the research community embraced biological ‘race’ in the NC genetic theory of ‘racial’ kidney disease and ignored medicontexts in subsequent research that pooled findings from around the US, **Fig 4**.

Moving kidney disease of Black Americans from “genetic” to “unknown cause” [91,92,93] and raising awareness of persistent segregation and structural racism might spur medicontextual hypothesis generation (i.e., from smaller areas) and broader data collection to explore SDOH and environmental toxins (see **Appendix: 5.6 Case study: North Carolina**).

### 3.1 Removing ‘race correction’

Our re-analysis found Black and White veterans share the same curve for KF, suggesting cofactors of veteran status minimized ‘racial’ disparities, **Fig 7**. Finding similar outcomes in veterans regardless of ‘race’ suggests non-genetic factors in kidney injury and should redirect emphasis from genetics. In this section, we consider alternative explanations for observed differences between racialized groups.

#### 3.1.1 Comparable conditions

##### (a) Screened volunteers

Implied (but typically unstated) in studies of US veterans are the educational and physical selection standards of the US Armed Services [94]. Long before veterans enrolled in research, servicemembers met written requirements that screen out most of the widespread US functional illiteracy [95], **Fig 13**, and physical requirements (including urine tests) that precede comparable access to military training, jobs, and healthcare. With rare exceptions, servicemembers earned access to Veterans Administration (VA) Healthcare by completing basic training, entering active duty, and serving honorably. Basic training (e.g., to avoid dehydration) may promote relevant habits in veterans.

**Fig 13.**
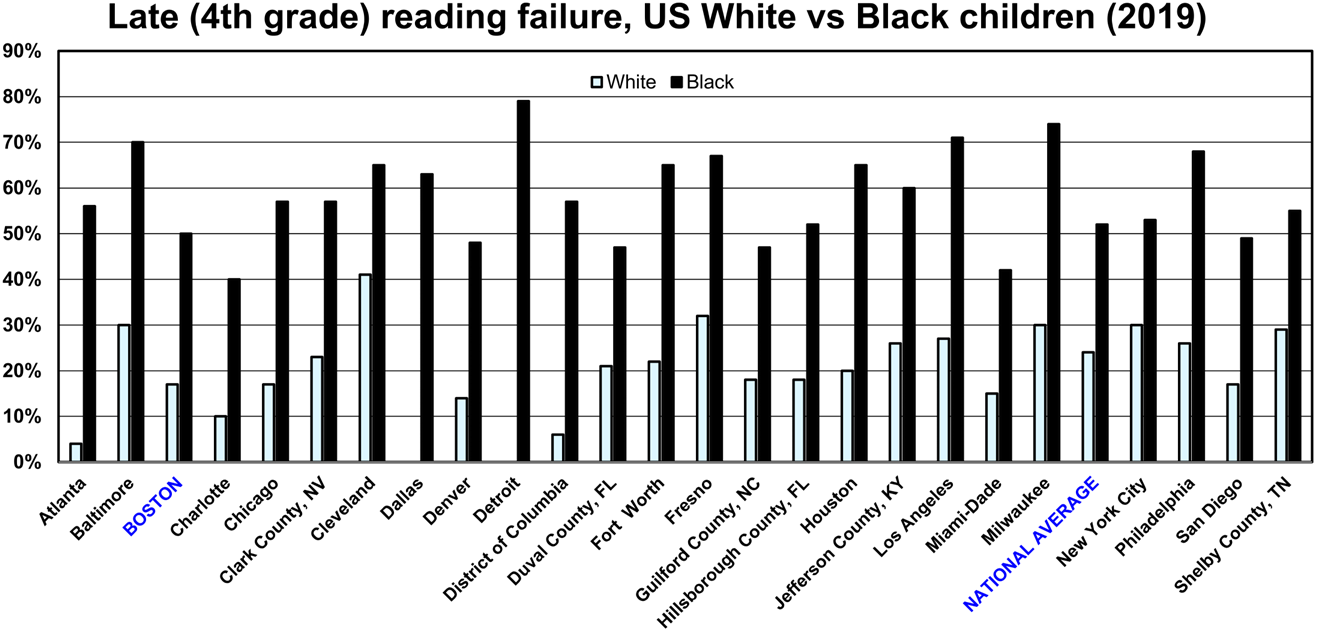
Late illiteracy: percentage of Black and White 4th grade students reading below basic level.

##### (b) Segregation

The European Commission noted that racial or ethnic discrimination “…can lead to a cycle of disadvantage which is frequently passed from one generation to the next” [7]. Peterson and Krivo demonstrated the relative advantage/disadvantage of US Black and White neighborhoods [96], **Fig 14**.

**Fig 14.**
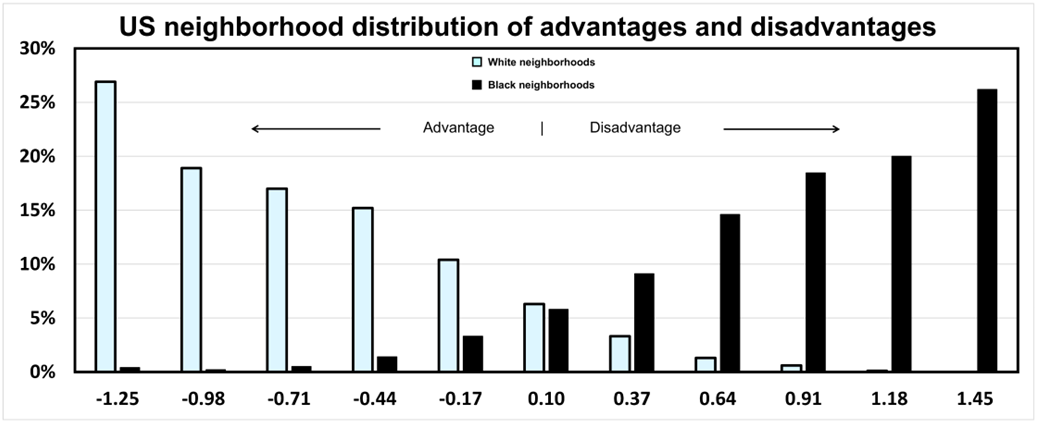
Distribution of advantages and disadvantages of US White and Black neighborhoods.

In contrast, US military communities are integrated, and Black service members fared better than Black civilians on numerous metrics [97]. Black and White veterans had similar employment rates [98] and more similar socioeconomic status, **Fig 5**. The actual economics of Black and White veterans might be closer because socially constructed ‘race’ influences residency options and choices. For example, Sampson and Bean noted, “…implicit bias in perceptions of crime and disorder may be one of the underappreciated causes of continued racial segregation” [99].

##### (c) ‘Racialized’ healthcare

Healthcare access is increased by higher income, having insurance, and higher education. In the general population, higher income was associated with lower CKD risk overall but higher CKD risk in *APOL1* HR individuals [100]. Similarly, stratifying the general population by self-identified ‘race’ showed “…higher income, having insurance, and higher education associated with a lower prevalence of CKD among White adults but not among Black adults” [101].

In the general population, it is intriguing that something associated with exposure to healthcare might, paradoxically, increase CKD risk in higher-income individuals with *APOL1* HR genotype and in Black adults with “higher income, having insurance, and higher education”. ‘Racialized’ prescribing in healthcare increases exposure to thiazide diuretics that can worsen dehydration. Initial treatment with “renoprotective” drugs might benefit all patients, including those with *APOL1* HR genotype [102,103].

##### (d) Environmental toxins

The “exposome” is the sum of pre- and post-natal exposures to environmental and socioeconomic factors [104,105,106,107,108,109]. Kidney injury is associated with a variety of environmental exposures, including drinking water contamination by heavy metals and volatile compounds [110,111,112], PFASs [113], agricultural toxins [114,115]. Even drought may affect kidneys [116]. Segregation is an intensifier of risk in the overall environment [117,118,119,120,121], health parameters [122,123,124,125], and access to healthcare and services [126,127,128]. Early exposures may “program” a path toward delayed kidney disease [129,130,131].

For example, there are disparities in lead exposure by ‘race’, education, and homeownership that vary by state and county [132,133], as shown, for example, in Detroit [134,135], Milwaukee [136], Philadelphia [137], and Washington DC [138]. Lead is associated with increased CKD risk, especially in youth [139,140,141,142]. Below age 5 years, lead exposure is far more prevalent in Black children [143]. Despite 90% reduction in US pediatric lead exposures since 1980, low-income and minority children in segregated urban schools and neighborhoods continue to have toxic lead exposures [144,145]. The acceptable threshold blood lead level (BLL) in children was reduced to 5 μg/dL [146], but some advocate zero tolerance [147]. Adults with a ‘normal’ blood lead level still may be more susceptible to kidney injury due to remote lead poisoning, especially adults born before recent lead abatement programs.

##### (e) Stereotyping

Black patients survive longer on dialysis. A study of dialysis survival after hospitalization for major medical illnesses blamed stereotyped personality traits, suggesting White patients give up on dialysis sooner than Black Americans with, supposedly, a greater sense of hope for the future from enduring hardships [148,149]. Unexplored alternative explanations include documented ‘racial’ differences in stroke mechanisms [150], dementia diagnosis [151], and other medicontextual processes, including racialized health [152]. Too early diagnosis of kidney failure and too early initiation of dialysis might follow ‘racialized’ use of thiazide diuretics in Black patients, potentially causing increased dehydration, serum creatinine, AKI, and CKD [49,50,51].

Stereotyping Black people (e.g., as ‘irrationally’ lacking trust in healthcare, lacking advance care planning [153], etc., [154]) is another example of victim-blaming [155,156]. Whether negative or positive, blaming the victim distracts from seeking medicontextual root causes. There is irony when the kettle pot of Blackness labels Black Americans as “lazy” [157]. Green noted, “…encouraging people to recognize information that is consistent with stereotypes may be helpful in dispelling damaging stereotypes within society” [158].

##### (f) Simpson’s paradox

One reason differences are often demonstrated in data is Simpson’s paradox, whereby results imply opposite findings based on whether estimates are stratified by key subgroups [159]. Until groups compared are adequately comparable, confounding remains a potential problem: it is impossible to attribute observed difference to one explanation or another because there are too many competing explanations for a fixed set of results. Simpson’s paradox is a particular risk when pooling medicontextual outcomes from multi-site studies [160,161], especially in the US, **Fig 4**.

Analyzing veterans separately from the general population may avoid Simpson’s paradox. Black veterans in VA healthcare had higher baseline comorbidities and illness severity yet equal or lower mortality to White veterans in all but six diseases [162] (or five, after our re-analysis). Despite a higher COVID-19 infection rate among minority women veterans, there were equal complications of mortality, cardiovascular events, or onset of heart disease among women veterans regardless of ‘race’ [163].

### 3.2 ‘Race’ and genetics

#### 3.2.1 Inconsistency of ‘race’

‘Race’, ‘mixed race’, ‘biracial’, ‘multiracial’, self-identified ‘race’—these terms are all bad science (BS) and invalid <u>inputs</u> for healthcare [16]. The medicontextual mechanism of action of socially constructed ‘race’ and *APOL1* genotypes in CKD must account for inconsistencies in folk taxonomies of ‘race’ [164,165,166,167]. For example:

***South Africa:***

> Evidence for race was found most familiarly in skin colour. But this was not necessarily the overriding or conclusive factor…. where they were born… had gone to school… where and with whom their children played… type and place of work… religious affiliation…. For example, a magistrate pronounced as Coloured a fair-skinned woman who was widely accepted as white…. prompted, it seems, by the fact that this woman had recently converted to Islam. [168]

***Brazil:***

> …there appears to be no racial descent rule operational in Brazil and it is possible for two siblings differing in Color to belong to completely diverse racial categories…. asked about their origins (the question admitted multiple responses) <10% of Brazilian black individuals gave Africa as one of their answers. [169,170]

***Canada:***

> Many different scientists, all of them white, undertook the complex work of delineating ‘racial’ categories Skin colour…. stature, head shape, cranial capacity, hair colour, hair texture, eye shape, eye colour, nasal index, and miscellaneous other facial features…. language, religion, geographic residence, manner of dress, diet, intelligence, reputation, and name were added to the list of identifying ‘racial’ elements…. [171]

***India:***

> Early attempts to classify race in India… relied on anthropometric measurements and often reinforced colonial biases…. categorized the Indian population into seven “physical types. …Colorism, with its preference for lighter skin tones, remains a pervasive issue, reflecting the enduring legacy of colonial influences and cultural norms. [172]

***Nigeria:***

> When I came to the United States, I hadn’t stayed very long, but I already knew that to be “black” was not a good thing in America, and so I didn’t want to be “black.” It took about a year of reading, learning, watching, for me to really come around and realize that there’s a context. [157]

US ‘racial’ classification originated in a 1662 statute [173]. The US medicontext is reflected in this segregation-era US Census instruction: “A person of mixed white and Negro blood should be returned as a Negro, no matter how small the percentage of Negro blood” [174,175].

In summary, the invention and tailoring of ‘race’ for local populations directly affects the medical literature. Medicontextualism accounts for authors and subjects in different countries absorbing conflicting local myths for coding ‘race’ (see **Appendix: 5.8 Black and “NOT Black”**). Electronic health records are often unclear who chose the label (i.e., actual patient self-report of personal lived experience versus healthcare provider assessment of skin color versus billing clerk [176]) and are unreliable [177]. The conflicting taxonomies cannot be collapsed into a single validated determination of ‘race’ (see **Section 3.3.3(c) Pharmaceuticals “harmonizing” ‘race’**). Thus, unstable, medicontextual ‘racial’ definitions and coding in large databases used for public health research produce unreliable findings that vary based on when, where, and by whom data was collected and articles written.

#### 3.2.2 Colorism and APOL1

“Colorism” varied the abuse of racism to stratify and stabilize all the colonial ‘racial’ hierarchies, correlating increasing pigment with worse deprivation:

> …the most effective way to segregate black people was to “use the dark skin slaves vs. the light skin slaves and the light skin slaves vs. the dark skin slaves.”.… Subsequently, colorism became an effective strategy in colonizing and dividing people. [178]

The odds of inheriting two, one, or zero copies of *APOL1* RV alleles vary, for example, with the number of great-grandparents of West African origin. With eight high-risk West African great-grandparents, all with *APOL1* HR genotype (i.e., two RV alleles), a subject will inherit two RV alleles. A subject with seven White great-grandparents and only one West African high-risk great-grandparent is considered Black in the US, “usually looks white or almost so” [179], and the odds of inheriting RV alleles are 75% for zero copies, 25% for one, and zero for two copies.

“In the US population, frequency of *APOL1* risk genotypes is estimated at approximately 15% in African Americans… and <0.01% in Europeans” [180]. In contrast, N264K**+** is more common in Europeans, found in less than 5% of Black American patients, and “…mutually exclusive with the *APOL1* G1 allele…. the presence of a single copy of the *APOL1* N264K mutation mitigated the increased risk conferred by HR *APOL1* genotypes” [181].

With *APOL1* RVs more common in West Africa and N264K**+** more common in Europe, people with one or the other regional distribution marker likely have different physical appearance and lived experiences. Thus, geographically localized “ancestry markers” can be proxies for ‘race’, and colorism can be a medicontextual mechanism stratifying subjects even in ostensibly ‘single-race’ studies.

#### 3.2.3 Confusing ‘race’, ethnicity, ancestry, and geography

Commonly used terms for socially constructed population groups are ethnicity in Europe and ‘race’ in the US. ‘Race’, ethnicity, and ancestry are medicontextually different (albeit similar and overlapping) concepts and all are subject to misuse.

NASEM recommended against conflating ‘race’ with ancestry:

> Conclusion 4-2. Using socially constructed groupings indiscriminately in human genetics research can be harmful. Their use reinforces the misconception that differences in social inequities or other factors are caused by innate biological differences and diverts attention from addressing the root causes of those social differences, which compromises the rigor and potential positive effect of the research. **Recommendation 1. Researchers should not use race as a proxy for human genetic variation. In particular, researchers should not assign genetic ancestry group labels to individuals or sets of individuals based on their race, whether self-identified or not.** [16]

Hung et al were not clear on what they meant by ‘race’ but expressed confidence in ‘race’ in their analytic approach to substantiate apparent relationships between ‘race’ and personal ancestry [74,78]. They described selecting a subset from the Million Veteran Program databank “…with genetic information available, of African ancestry” and used the method of “genetically inferred ancestry” described by Fang et al, in 2019 [182]. Their approach used principle components analysis to identify people with similar genetic profiles, which also tended to show overlap with racialized groups.

NASEM subsequently noted:

> There is a pervasive misconception and belief that humans can be grouped into discrete innate categories…. The illusion of discontinuity between racialized groups has supported a history of typological and hierarchical thinking…. These modes of thinking often spill over to other descent-associated population descriptors such as ethnicity and ancestry (Byeon et al., 2021; Fang et al., 2019). The structure of human genetic variation, though, is the result of human population movement and mixing and so is more related to geography than to any racial or ethnic classification. [16]

People who look similar are more likely to share ancestry and genetic overlap. The problem is ignoring arbitrary injury attracted by ‘race’ in the medicontexts derived from colonialism, slavery, and global migration. Genetics cannot be separated from this confound. ‘Race’ ethnicity, and ancestral history predict medicontextual differences in many outcomes. Careful research might distinguish effects of social treatment and generational wealth in daily life. To presume the medicontextual mechanism of difference is genetic is bad science (BS).

#### 3.2.4 ‘Race’ in research design

NASEM recommended broader and more detailed investigations:

> Conclusion 4-8. In the absence of measured environmental factors, researchers often wrongly attribute unexplained phenotypic variance between populations to unmeasured genetic differences. **Recommendation 4. Researchers conducting human genetics studies should directly evaluate the environmental factors or exposures that are of potential relevance to their studies, rather than rely on population descriptors as proxies.** [16]

The Hung et al data are valuable but the ‘single-race’ study design presupposed an association between genetic biology and medicontextual ‘race’, made analytic assumptions overlooking more direct causes of AKI, whether or not captured in data, and gave the appearance of *APOL1* genotypes justifying the myth of ‘race’ as biology [74,78].

The directed acyclic graph (DAG) [183] in **Fig 15** is a causal framework modeling their choices, but with modifications showing potential “back door paths” that explain the medicontextual outcomes better than a direct effect of APOL1, including possible differential effects of medicontextual racism between *APOL1* HR and LR genotypes. The grey oval represents their focus on biology. The rest of **Fig 15** shows the larger medicontext that our re-analysis of veterans suggests is important. Their meta-analysis models included some demographic variables, but none related to education, employment, colorism, socioeconomic status, or any other medicontextual SDOH. Nor did they justify choosing <u>an</u>y variables for the analysis: adding more to a model can introduce selection bias by way of Simpson’s paradox.

**Fig 15.**
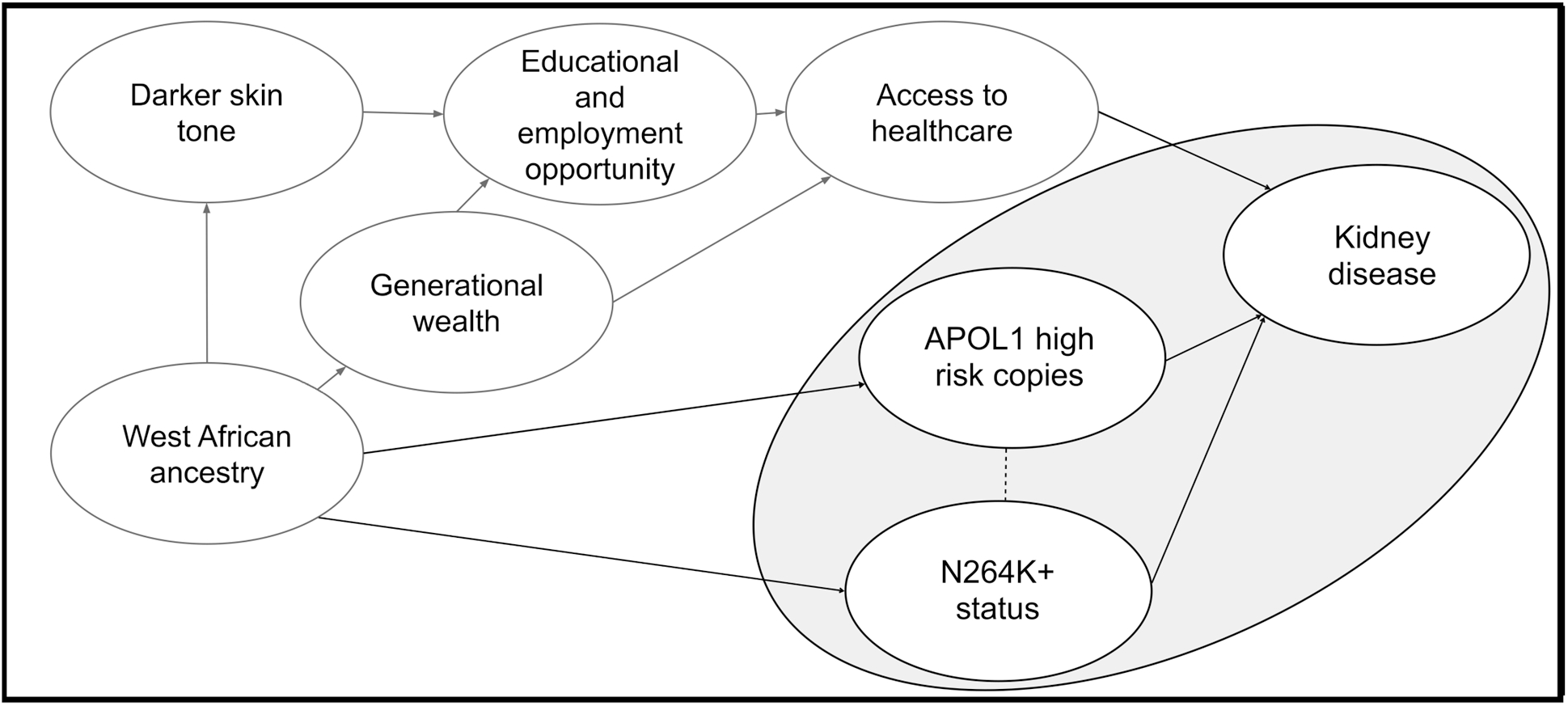
Causal directed acyclic graph (DAG) that the study might have imagined (with our modifications). One back door path from *APOL1* to kidney disease is via West African ancestry (i.e., West African ancestry increases odds a person has *APOL1* high-risk copies but also alters physical appearance, and physical appearance feeds discrimination). The study assumed the 10 ancestry principal components (plus other variables in other models) analytically controlled for this to justify focusing on relationships in the shaded area. First, we disagree that ancestry principal components controlled for the back door path via skin tone. Second, even if principal components addressed the confound of skin tone, that does not account for the history of discrimination that is its own confound, including generational wealth as a significant factor. Third, given their regional dependence and low frequencies, the inclusion of both N264K and *APOL1* in one analysis is problematic. This approach leads to redundancy and collinearity, resulting in uninterpretable coefficients.

We found several potential back-door paths that the research design did not address. First, physical appearance: West African ancestry, darker skin tone, and medicontextual colorism. Second, generational wealth: even Black Americans who could ‘pass’ as White had medicontextually limited opportunities that denied generational wealth. Third, the mechanism of N264K “protection”: the low frequency of N264K variant in African-American veterans (4.3%) likely also correlates with physical traits. Further, we do not know whether all haplotype combinations truly exist in nature, and a more plausible explanation for the infrequent genetic combination in this sample is that they are simply false positives rather than representing something meaningful.

#### 3.2.5 Equity and APOL1

**Ethicist Dorothy E. Roberts** noted, “The issue is not whether genes affect health—of course they do—but whether *genetic difference* explains *racial disparities* in health” [184].

The APOL1 mechanism that explains medicontextual ‘racial’ differences in KF is unknown [185]. Higher RVs of the *APOL1* gene were identified in all haplotype backgrounds, with magnitude of effect dependent on expression levels and equity a confounding factor in these findings. Because overexpression of <u>any</u> form of APOL1 is cytotoxic, *APOL1* variants appear non-significant [186].

‘Racial’ medicontextualism notes multiple mechanisms wherein racism affects COVID outcomes. These include misdiagnosed lung disease [187,188,189], mismeasured blood oxygen levels [190,191], and inequitable care [192,193,194]. The different ‘racial’ systems in Brazil and the US converge in colorism, but medicontextual links between ‘race’, skin color, and ancestry varied under different ‘racial’ systems. A broader range of physical appearances and SDOH might weaken *APOL1* associations with ‘race’ in the Brazilian population (i.e., more noise in the data) [195]. Given the wide confidence intervals in the Brazilian data [77] and a general caution with accepting the null hypothesis of no difference, we cannot say the ORs are significantly different when comparing the same risk factors between the two samples, **Fig 10**. However, the remarkably similar ORs between US veterans and Brazilians for a variety of non-genetic covariates support other aspects of personal medicontext as more important than the *APOL1* gene [196,197]. Similar to veterans in VA Healthcare, Brazilians access universal healthcare as a constitutional right, and although some buy private supplemental insurance, there was no difference between “white and non-white” Brazilians who were Dual Users of public and private healthcare services vs. Non-Dual Users [198].

Skidmore noted,

> Brazilian society was based on an explicit belief in white superiority, although not white supremacy. The distinction is crucial…. white stood at the apex of the social scale and African at the bottom. But the Portuguese in Brazil differed from the English in North America. [199]

Multiple US general population studies of *APOL1* genotype found that adjusting for some combination of age, sex, baseline eGFR, income, and education minimized or eliminated correlation between KF, *APOL1*, and ‘race’ [200,201,202,203].

#### 3.2.6 Focal segmental glomerulosclerosis

Our re-analyses did not conclude that *APOL1* HR genotype has no association with AKI and CKD. Rather, the association appears overstated. Hung et al showed *APOL1* HR genotype associated with modestly increased risk of FSGS in US veterans [78], consistent with other recent reports [204,205,206,207], including low penetrance [208]. However, FSGS is nonspecific and may reflect multiple medicontextual cofactors that veteran status mitigated to relatively few FSGS and KF events, small absolute percentage rates, and ORs within the block of modest ORs, **Fig 10** and **Fig 11**.

##### Second or first hits

Descriptions of FSGS histology on light microscopy range from “minimal change” to collapsing glomerulopathy (CG)—a form of podocyte injury wherein, some say, “Even a single glomerulus with these histologic features… is sufficient to diagnose CG” [209]. Classifying FSGS by etiology [210,211] defines four subcategories represented in **Fig 16** as thick-bordered rectangles: primary, secondary, genetic, and unknown. Two paths show competing classifications for Black patients, both starting from the pink diamond in the upper left and ending at the rectangle in the lower right.

**Fig 16.**
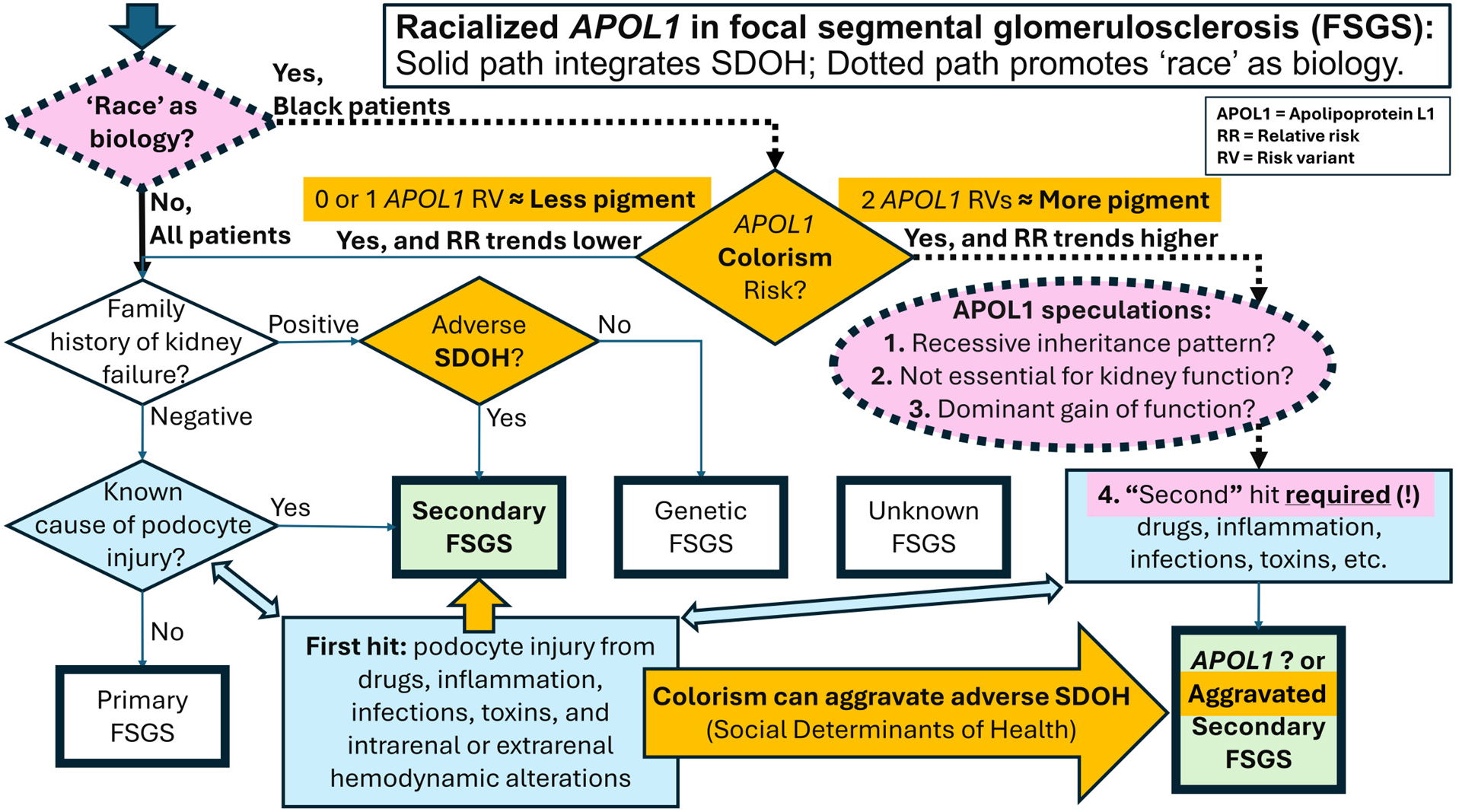
‘Racialized’ APOL1 in FSGS classification. Some promoted correlation between *APOL1* and ‘race’ as “proof” that biological ‘race’ underlies ‘racial’ disparity in kidney failure (KF). APOL1 mechanism is unknown and not essential for kidney function. Characteristics of “*APOL1*” FSGS that are atypical for genetic kidney diseases brought strained speculations for a unique classification. Increased family history of KF across *APOL1* RV genotypes suggests *APOL1*-linked genes and SDOH contribute to familial clustering (see text). Structural racism (e.g., colorism on pigment gradient correlated with number of *APOL1* RVs) aggravates adverse social determinants of health (SDOH). “*APOL1*” FSGS is just aggravated secondary FSGS. The dotted path, misusing ‘race’, might then be misused to justify “APOL1” drugs to Black patients.

##### Solid path: Environment and SDOH

The solid path, down the left side and across the bottom, begins with family history and presents ‘race’ as attracting socially constructed injuries. CKD with a family history of KF often reflects common environmental exposures, regardless of *APOL1* genotype [212]. Known causes of secondary FSGS include drug exposures, infections (viral and other), autoimmune diseases, malignancies, ischemia-related conditions, genetic diseases, and “Superimposed on other kidney diseases” [213]. Along this “solid path”, “second” hits are the etiologies for “secondary FSGS” in all patients (e.g., HIV [214] and small particle air pollution [215,216]), with environmental and SDOH injuries aggravating secondary FSGS (represented in **Fig 16** by the rectangle at the lower right), especially Black patients with *APOL1* HR genotype and more of the pigment targeted by colorism.

##### Dotted path: Biological ‘race’

The dotted path, across the top and down the right side, promotes biological ‘race’. *APOL1* HR genotype is so unlike other genetic causes, it occupies a separate category (also represented in **Fig 16** by the rectangle at the lower right). The dotted pink oval lists speculations to explain *APOL1* atypicality, including **(1)** recessive-inheritance pattern that usually signals loss of an essential function. However, **(2)** apolipoprotein L1 is not essential for normal kidney function [217], so **(3)** *APOL1* RVs would have to represent gain-of-function mutations, but those usually shows a pattern of dominant inheritance. Because only about 20% of patients with *APOL1* HR genotype develop kidney disease [218], which is atypical for a dominant disease, speculation hypothesized **(4)** the need for a second hit” [219]. On the dotted path, Black Americans <u>without</u> *APOL1* HR genotype revert to the solid path and sometimes end with typical secondary FSGS. Black Americans <u>with</u> *APOL1* HR genotype follow the dotted path to required “second hits” (strikingly similar to those for typical secondary FSGS [213, 220]) and may end with “APOL1 FSGS”. Researchers promoting biologic ‘race’ seem to claim a distinction between “APOL1 FSGS” and secondary FSGS aggravated by colorism.

##### FSGS confusion

**Alexander J. Howie, Professor of Renal Pathology at University College London**, described confusion in reports of FSGS:

> FSGS has been applied to such a wide range of conditions that it can reasonably be said to have no useful meaning. Attempts to refine the term have been largely ignored. Study of 252 papers on genetic causes of FSGS found various clinical features. Many papers took the reported diagnosis without question. Few papers reported a pathological review, almost half reported FSGS and up to six other conditions caused by any particular gene, some reported FSGS with recognisable glomerular disorders, over 80% did not apply the Columbia classification, and in nearly all with photomicrographs, the images were not useful for refinement of FSGS. Some workers commented on a lack of genotype-phenotype correlation….
>
> These findings on the selection, diagnosis, description and illustration of cases show that the literature on genetic causes of FSGS is in confusion…. At the moment, there can be little certainty that any paper on FSGS is studying the same condition as any other paper…. [221]

Blaming CG on genes (e.g., *MHY9*, *APOL1*) overlooked its absence prior to and increasing prevalence since the late 1970s, suggesting a new etiology. Example candidates include new environmental exposures (e.g., new food additives or prescribed medicines) or policy changes to SDOH (e.g., increased access to healthcare after the Civil Rights Movement [222]). Coincidentally, earliest reported CG roughly coincided with increased treatment of hypertension [223]. Goal-oriented BP therapy from the late 1970s [224,50] increases AKI in Black patients [51], who are more often dehydrated [42,43] and ‘racialized’ with thiazide diuretics [47,48,44,45,46].

##### Global origins

Stereotyping dark skin as a sign of *APOL1* HR genotype follows enslavement of West African people, with their descendants still living in the US or recently immigrated from Caribbean countries. However, more recent US immigration, fueled by conflicts in Central and East African countries, continues to diversify African ancestral histories. Even if visual examination increased the likelihood for *APOL1* HR genotype, who and when to test and counsel [225] hinges on validity of the solid versus dotted paths, in **Fig 16**, and understanding reduced disparity in veterans. Nonspecific Population Health measures may mitigate accumulated ‘second hit(s)’ from SDOH, even before any component cofactors are fully characterized.

### 3.3 Potential implications

In *Demolishing Detroit: How Structural Racism Endures*, Caverly noted:

> Racist outcomes without racist intent are the calling cards of structural racism. In the United States and elsewhere, racial disparities in experiences and life chances continue to be relatively unchanged long after racist ideologies and policies have been seemingly rethought. [226]

Medicontextualism attends to the idiosyncratic mechanisms and consequences of structural racism under different economic systems. Deprivation relative to the dominant group is universal and is associated with CKD [227,228]. Other components, like exposure to specific toxins, vary with local economic activity, such as the level of mining, industrialization, and sanitation. The success of managing exposures varies with local availability, quality, and racialization of healthcare and education. Because economic activity may vary greatly over short distances, medicontextual cofactors can show marked local heterogeneity, especially in larger nations (e.g., **Fig 4**).

Thus far, we have focused on bad science (BS) in the medical literature that can mislead clinical practice by misattributing the mechanisms for medicontextual differences between racialized groups. The implications of this misattribution extend beyond academics and basic clinical practice. The much larger concern is how a presumed biological basis for ‘race’ shapes medical research and the development of new treatments. The advice of **John P.A. Ioannidis, Professor of Medicine and Epidemiology at Stanford University**, is relevant:

> Before running an experiment, investigators should consider what they believe the chances are that they are testing a true rather than a non-true relationship….
>
> Corollary 4: The greater the flexibility in designs, definitions, outcomes, and analytical modes in a scientific field, the less likely the research findings are to be true….
>
> Corollary 5: The greater the financial and other interests and prejudices in a scientific field, the less likely the research findings are to be true. [229]

Commercial interests drive innovation, but there is also profit potential in misuse of ‘race’ [230]. Research into low-cost preventive measures and medicontextual SDOH usually requires government funding, so we highlight three such areas for further study: **(1)** US illiteracy and deprivation cofactors implied by outcome differences between veterans and the general population (see **Section 5.5 Bias in FSGS and APOL1** and **Section 5.6 Case study: North Carolina** for some medicontextual cofactors), **(2)** role of ‘race’ when searching for genetic causes without a medicontextual hypothesis, and **(3)** international collaboration on ethics that could discourage misuse of ‘race’.

#### 3.3.1 Proxy for illiteracy and deprivation

##### (a) Illiteracy

A 2001 summary of decades of reading research funded by the National Institutes of Health (NIH) concluded:

> Failure to develop basic reading skills by age nine predicts a lifetime of illiteracy…. On the other hand, … provision of comprehensive early reading interventions can reduce the percentage of children reading below the basic level in the fourth grade… to six percent or less. [231,232]

Illiteracy varies by US district, **Fig 13**, and may mediate the medicontextual relationship between US ‘race’ and CKD [233,234], implicating broad consequences of reading failure (e.g., difficulty obtaining jobs with US health benefits that are not universal). Up to 90% of US high school g<u>raduates</u> may be illiterate [235]. Further, lead exposure affects educational outcomes [236,237,238,239,240], which may contribute to difficulty remediating reading failure.

Kozol observed,

> …education is not a protected right under the US Constitution.… [P]roperty-rich districts in areas surrounding our cities are almost all predominantly white…. [T]he near total racial isolation of children of color in our nation’s cities is the present-day reality of schooling that is separate and unequal. [241]

In contrast, literacy diminished the association between long-term illness and Black ‘race’ and removed the predictive power of “education” and being African American [242]. Adult literacy programs did not improve CKD [243], but <u>early</u> intervention can succeed [244,245,246,247]. Districts incorporating NIH reading research [248] (e.g., through high-quality programs like LETRS [249]) have shown progress (e.g., Bethlehem, PA [244], Easton, PA [246], Mississippi [250,251]). Universities could ensure that their Schools of Education incorporate NIH reading research into teacher training [252,253,254].

##### (b) Deprivation

The origin of US housing segregation “…has now largely been forgotten” [255,256]. However, patients from disadvantaged neighborhoods [257] continue to suffer health effects of childhood poisonings and prenatal exposures [258,259] that increase CKD risk [260,261,262]. Novel approaches to reducing lead exposure [263] have low profit potential and delayed benefit due to a lag between prenatal or childhood lead exposure and long-term consequences [264].

Early detection of pre-chronic kidney disease (preCKD) [1] could allow early intervention [265]. Further research (e.g., in zip codes of the few White and Black neighborhoods with similar demographics, **Fig 14**) might show whether early toxic exposures [266] contribute to medicontextual ‘racial’ disparities in GFR and to the purported “senile” nephrosclerosis found in one-third of young but absent in 10% of elderly kidney donors [267]. Examining data on KF by zip code [73] may show an effect of veteran status on local ‘racial’ disparities [29].

Deprivation is “…more powerfully associated with brain function and structure than other variables… [E]nvironmental factors indexed by neighborhood socioeconomic status, including sleep and stress, strongly shape childhood brain organization” [268]. Literacy testing <u>alone</u> [269,270] could prevent medicontextual ‘race’ from being a proxy for illiteracy.

#### 3.3.2 Racism’s genome-wide associations

Genome-wide association studies (GWASs) often search outcomes data without a medicontextual hypothesis and often without a known mechanism of action. The goal often is to identify genes relevant to disease development, but there usually is not a particular gene in mind prior to implementing the analysis. A GWAS promoting *APOL1* suggested, “…the genetic risk that was previously attributed to *MYH9* may reside, in part or in whole, in *APOL1*, although more complex models of risk cannot be excluded” [271]. “More complex models” includes medicontextual stratification of resources by skin pigment—structural racism—which has clear medicontextual mechanisms of action.

Kidney disease mortality rates vary more by US city than between some countries with ‘corrections’ for nationality, **Fig 4**. This might suggest the need for research into city-specific *APOL1* gene frequencies, but our findings in veterans support modifiable environmental and social cofactors, suggesting *APOL1* may follow *MYH9* into irrelevance (see **Appendix: 5.5 Bias in FSGS and APOL1**). Simpson’s paradox appears likely when misusing ‘race’ and pooling cohorts, limiting cross-medicontextual utility of polygenic risk scores (PRSs) for kidney disease [180,272,273,274,275].

Colorism enhances the illusion of ‘race’ as biology because social injuries often vary with polygenic pigment, both directly (e.g., through prenatal and childhood health, education, employment, access) and indirectly (e.g., through circumstances of parents and grandparents), especially confounding GWASs [276]. A ‘structural racism’ polygenic risk score (srPRS) based on a subset of 160 pigmentary and other racialized genes [277,278] would likely capture colorism’s gradient of socially-constructed (i.e., ‘racial’) insults. An srPRS might help researchers to perceive that racism indeed works through genes, and help to end circular reasoning from racism to theories of biological ‘race’ to single-nucleotide polymorphisms (SNPs) in linkage disequilibrium with racialized genes (i.e., the srPRS components).

Pigment has physiologic properties (e.g., on pulse oximetry [190,191]) that can affect results but are not ‘racial’ (i.e., will not apply to all or only Black people). Researchers who select subjects, consciously or unconsciously, based on appearance or socioeconomics may unwittingly skew the effects, for example, by testing an optical device only in White people (i.e., the residual exclusion of structural racism) or selecting a cohort of Black Americans with the most pigment and, typically, the least resources due to historically harsher experiences under colorism (see **Section 3.2 ‘Race’ and genetics**). Further, associations alone cannot tell whether historical violence against groups of people based on appearance induced heritable epigenetic changes in gene function—interactions between medicontextual social environment and gene expression.

Instead of combining Black veteran and Black general populations [279,159], GWASs comparing their results might reveal SNPs that appear under racialized socioeconomic and gene-environment stresses (e.g., illiteracy, deprivation).

#### 3.3.3 Appeal to bioethics

Because ‘race’ is socially constructed, our research merely confirms well known facts: conclusions of <u>biological</u> ‘race’ will always be false, and re-analyses will always find social causes (medicontextualism seeks the “Where’s Waldo” ‘racial’ assumptions in articles that stop at ‘race’ as biology). Research providing no explanation for ‘racial’ differences or no medicontextual justification for including ‘race’ beyond “predictive accuracy” is especially problematic. As NASEM noted:

> …studies that are poorly designed to answer research questions are scientifically invalid and unethical…. What is necessary is an understanding of the underlying issues during study design and long before data analyses: the moment of publication is far too late. [16]

If ethical oversight or journal editors or grant funders required, at the least, broad measures to capture the medicontext of research studies (i.e., SDOH, including for housing, education, toxic exposures, deprivations, etc.), Jones noted, “…researchers would fall in line immediately…. [T]hen we would have much better datasets, and it would be possible to figure out what is this race signal coming from?” [280].

We briefly discuss legacy commitments that quietly sustain misuse of ‘race’ through **(a)** data, **(b)** medical journal traditions, and **(c)** international regulatory agreements. We also advocate **(d)** trust through institutional review, which appears crucial to ending misuse of ‘race’ as an input to healthcare.

##### (a) Legacy data from pre-ethical ‘race’

###### Retroactive manipulation

In 1999, MDRD Kidney researchers proposed the first ‘race corrected’ eGFR [281]. Their demographic “Form # 04” [282], **Fig 17**, shows “Hispanic” under ‘race’ **(left)** and no instructions for assigning “Race/Population Group” under question 10 **(right)**. In 2006, MDRD authors confirmed the omission: “Ethnicity was assigned by study personnel, without explicit criteria” [283]. In 2021, 22 years after the first study’s moment of publication, an unsigned “correction” claimed MDRD used “self-identified race” [284] (referencing “Form # 03” [285] that lacks mention of ‘race’). NIH separated ‘race’ from ethnicity and adopted self-identified ‘race’ 11 years <u>after</u> MDRD Form # 04 was last revised [286]. Burke and Corwin commented, “…transition to self-identification of ‘race’ would have been *far* ahead of those times and would have required a remarkable degree of empathy, study-related re-education, and… a paper trail” [287] (the comment appeared below the “correction” but was never indexed in PubMed). Lack of response to this timely comment suggests the medical journal editors may not know the identity of the author(s) of the “correction” or that no ethical standards compelled release of the name(s) of the author(s), response to the charge of “faulty recall”, or public debate before altering a 15-year-old text. The change likely prolonged citations to, and clinical use of MDRD eGFR despite <u>then known</u> risks for Black patients: “When applied to cohorts outside the original study, both the MDRD and CKD-EPI equations have proved to be highly biased, imprecise, and inaccurate when compared to measured GFR” [288].

**Fig 17.**
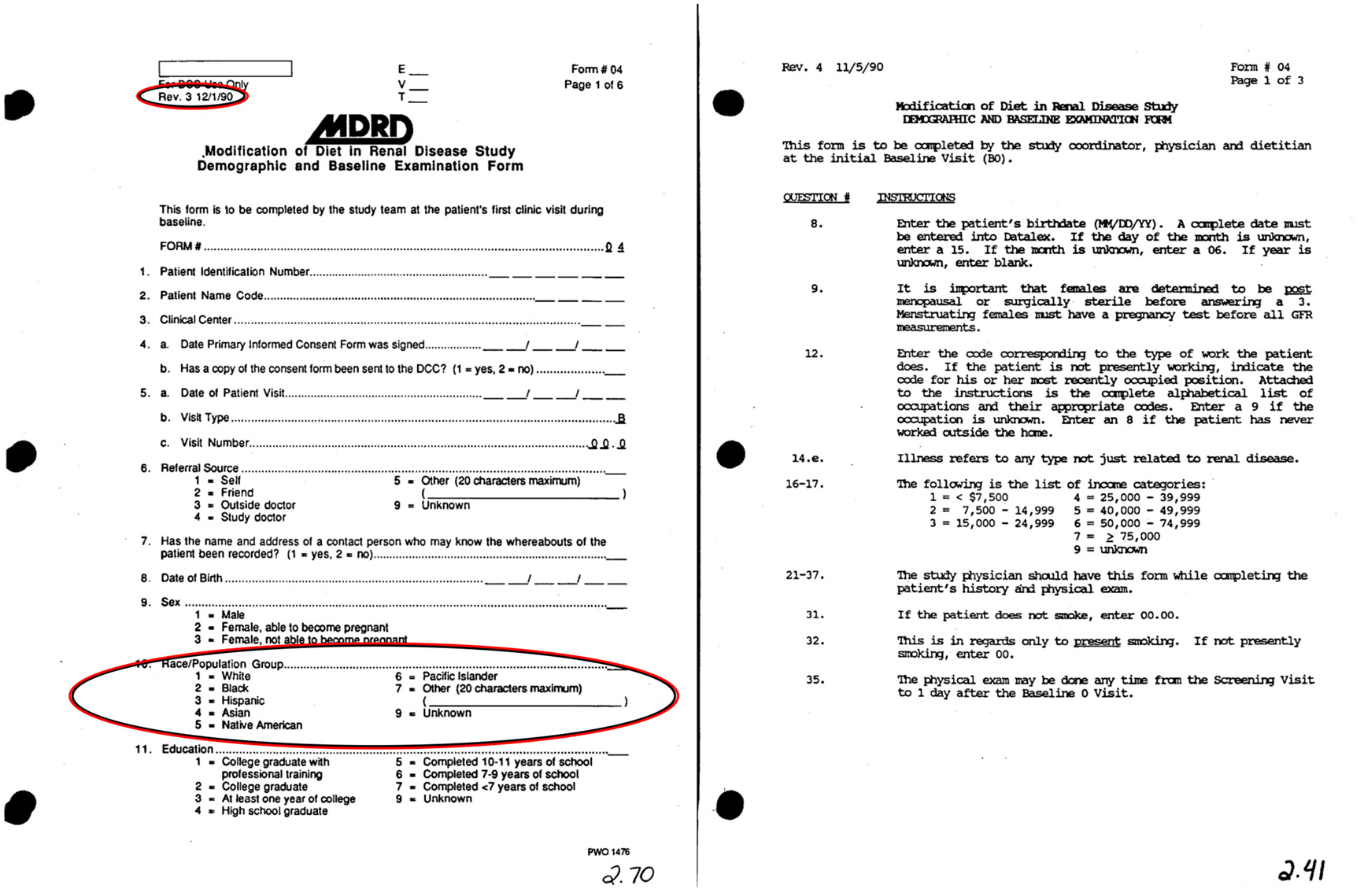
MDRD Study Form 04 - Demographic & Baseline Exam case report form template (left) and instructions (right). Revised on “12/1/90” (upper left), the “Race/Population Group” (bottom left) is consistent with 1990s requirements. The lack of instruction for ‘race’ (right, question 10) is inconsistent with an early shift to self-identified ‘race’. NIH separated ‘race’ from ethnicity and adopted self-identified ‘race’ in 2001, “EFFECTIVE… January 10, 2002”.

###### Aware without care

Bad science (BS) studies can perpetuate the ‘race’ myth in ways that are “largely unintentional” [289], but in 2008, **HUNT** and Megyesi found that most genetics researchers **(1)** understood that ‘race’ is invalid, **(2)** used ‘race’ as a proxy for biological cofactors without bothering to explore further (simply assumed cofactors would be difficult to identify and measure), **(3)** could list serious potential negative consequences of misusing ‘race’, but **(4)** did so anyway [290]. After the NASEM report [16], printing the same article in five journals [291,292,293,294,295], genetics journal editors encouraged the allowed form of social commentary: genetic scientists rationalizing legacy misuse of socially constructed ‘race’ to continue publishing bad science (BS) under “…the tranquilizing drug of gradualism” [296]. Their predecessors did the same, 75 years ago:

> For the social scientists gathered by UNESCO to write its 1950 ‘Statement against race and racial prejudice’, race had to be disproved on scientific grounds, a task which they saw as easily achievable. The Statement claimed that:

> The division of the human species into ‘races’ is partly conventional and partly arbitrary and does not imply any hierarchy whatsoever. Many anthropologists stress the importance of human variation, but believe that ‘racial’ divisions have limited scientific interest and may even carry the risk of inviting abusive generalization. (UNESCO, 1968: 270)

> Directly after the publication of the Statement, it was followed by an alternative Statement written mainly by geneticists also involved in the UNESCO project…. that race continued to be of scientific usefulness for describing ‘groups of mankind possessing well-developed and primarily heritable physical differences from other groups’ (Comas, 1961: 304) and should not be confused with politics. [297]

###### In 2025, Jerome P. Kassirer, a nephrologist and former Editor-in-Chief of The New England Journal of Medicine, (NEJM) and Robert Steinbrook, a former editor at NEJM, wrote

> In 1980, Dr. Arnold S. Relman, then the editor-in-chief of the New England Journal of Medicine, famously warned about “the medical-industrial complex,” a “huge new industry that supplies health-care services for profit,” and the necessity of ensuring that the interests of the public come before those of stockholders. Nearly 50 years later, in the era of private equity investment in health care, that warning has never been more prescient….
>
> Standing up for patients… is public service, not a partisan political activity. [298]

###### Healthcare or research

**Medical ethicist Evelyne Shuster** described “…the continuing difficulty physician–researchers have with accepting public regulation of research” and the ethical lapses of US doctors testifying at the Nuremberg Medical Trial that inspired

> …moral rules centered on the participant and not on the physician….
>
> Ivy repeatedly said that he relied on Hippocratic ethics, which govern the doctor–patient relationship. In the end, however, he folded, undermining his entire testimony…. He had to admit that there were differences between a treating physician who acts in his patients’ best interests and a physician–investigator whose goal is to test a scientific hypothesis for the benefit of society rather than the participant. [299]

Research governance “for the benefit of society” [300,301] has allowed misuse of the contributions of Black participants to worsen healthcare for Black patients [38,66]. **Medical ethicist Harriet A. Washington** documented ongoing ethical lapses of medical research in *Medical Apartheid: The Dark History of Medical Experimentation on Black Americans from Colonial Times to the Present* [302]:

> Today, the worst abuses are mostly memories, although some forms of abusive research persist, and a few new ones have arisen…. The long history of flawed science in the service of preconceived notions is being supplemented by new, insufficiently questioned racial theories of disease. Adopting these unquestioningly while ignoring important environmental disease factors not only imperils black health; it also reinforces the idea of blacks as possessing dramatic physiologic differences. [302]

##### (b) Legacy traditions of US medical publishing

The origins of US medical publishing offer insights. The American Medical Association (AMA) described the determination of its founder “…to explicitly exclude women and Black physicians…. [His] role was highly active, not passive, and his choice for a racist direction was pursued with energy and force” [303,304].

In 1880, as founding editor of *The Journal of the American Medical Association* (JAMA, “…the most important US journal in the first half of the 20th century” [305]), this racist man established the traditions of bad science (BS) in medical publishing that keep medical research inside the grey oval, **Fig 15**, blocking study of the role of ‘race’ in research findings. Like other journals that rejected our claim of relevance (e.g., to discuss the medicontext of illiteracy and sociology of deprivation in our research), JAMA is known for excluding discussion of the racism behind socially constructed ‘race’ and for segregating consideration of social cofactors in articles labeled “Commentary” and “Perspective” [2].

The AMA persistently opposed universal healthcare [306]. The federal government linked health insurance to employment but excluded agricultural and domestic workers [307,308]. Without mentioning ‘race’, these policies denied healthcare especially to Black people, and others, with broad consequences for the research medicontext [309].

Judging past actions and events for adherence to the ethics of their time is the domain of historical ethics. Recounting the antebellum ‘racial’ theorizing of a founder of *The New England Journal of Medicine*, **Historical ethicists Christopher D.E. Willoughby, Dominic Hall, Evelynn M. Hammonds, and Scott H. Podolsky** et al noted:

> Leading journals published debates over whether Black people and White people were different species. As they built enduring U.S. medical institutions, largely improving medical knowledge and practice and standardizing training, they wove racist theories and norms into the fabric of American medicine.
>
> Understanding racism in American medicine requires scrutinizing not only the most extreme beliefs and behaviors but also everyday practices and routines. For centuries, physicians normalized racist ideas…. Reversing this long historical process requires more than just reaction or reckoning: it will require complex work to fully disentangle racial thinking from medicine [310,311,312].

Editors with different lived experiences of ‘race’ could enhance “peer review” of medical research, but decent, well-meaning colleagues who edit medical journals are overwhelmingly “not Black” [313]. Blind spots allowed misuse of ‘race’ in the articles we noted above, suggesting the philosophy of human medical science needs reexamination [314,315,316,317,318]. And because advocating change takes more words than “stay the course”, modern word, figure, and reference limits also favor the status quo (i.e., “…the master’s tools will never dismantle the master’s house” [319]).

##### (c) Pharmaceuticals “harmonizing” ‘race’

‘Race’ persists through active institutional maintenance [320]. The Minority Health and Health Disparities Research and Education Act of 2000 required collection of ‘racial’ data that facilitated the “pharmaceuticalization of race” [321]. **Medical ethicists Shai Mulinari, Andreas Vilhelmsson, Piotr Ozieranski, and Anna Bredström** found sharply increased ‘race’ and ethnicity data on drug labels, encouraging ‘racialized’ perceptions in medical care:

> …in 2004–2007, 46% (37/81) of US labels contained race/ethnicity demographic information… versus 80% (171/213) in our 2014–2018 sample… the routine report of race/ethnicity demographics and analyses may prime prescribers to believe that race and ethnicity are biologically and therapeutically meaningful *even* when labels suggest no differences and particularly when they assert minor or ambiguous differences. [322]

**Medical ethicist Jonathan Kahn** described the marketing utility in misuse of ‘race’:

> Medical researchers may say they are using race as a surrogate to target biology in drug development, but corporations are using biology as a surrogate to target race in drug marketing…. It is far easier to target African Americans than to identify a market of particular individuals who happen to respond well to BiDil because of their genetic makeup regardless of race. [230]

The EU rejects use of ‘race’ at home [7] but lends international credibility to US promotion of <u>biological</u> ‘race’ [323]. **Ethicist Mulinari** et al summarized the history:

> …much empirical and theoretical work has focused on BiDil, but the European Medicines Agency (EMA) never approved this drug….
>
> …the ICH-E5 document, “Guidance on Ethnic Factors in the Acceptability of Foreign Clinical Data” …. was developed by regulators and industry in the EU, the USA, and Japan… to harmonize requirements for drug development and regulation under the auspices of the International Conference on Harmonization (ICH)…. The EU did not want to “overemphasize diversity” between races; the USA… “hoped [that] the US [racial and ethnic] standards would be accepted as universal”; and Japan emphasized that… the Japanese could not be subsumed under a diverse “Asian race,” …. Despite the reported little EU interest and opposition from Japan, the final ICH-E5 document suggested… three “major racial groups,” i.e., Asians, Blacks, and Caucasians. [322]

Noting the ICH process, **ethicists Mulinari and Bredström** summarized, “Thus, ethnic sensitivity is due to ‘ethnic factors’… either ‘intrinsic’ or ‘extrinsic’, and in the ICH definition, race is solely ‘intrinsic’, and more specifically *genetic*” [324]. **Ethicist Kahn** also noted,

> …in a world rife with historical examples of stigma and discrimination based on false constructions of particular groups as biologically different (and/or inferior) the geneticization of race is fraught with peril. One need not identify specific, immediate consequences that might flow directly from the ICH guidelines to caution that the formal, regulatory adoption of genetic conceptions of race could contribute to unintended and unforeseen negative consequences. [325]

“Self-identification” of ‘race’ projects a veneer of legitimacy, but victims of shared hardships of racism and ethnocentrism must “choose” their label from a constrained, globally “harmonized” list curated by the US Food and Drug Administration (FDA). **Ethicists Mulinari and Bredström** noted “collaborative” international regulation that reinforces US ‘racial’ categories and “…endorses the use of the FDA-requested racial categories… regulatory and academic science that stabilise the idea of race as a key biomedical variable” [326]. And **ethicist Washington** described pharmaceutical industry infiltration of medical journals for the exploitation of ‘racial’ categories and “…the much-abused technique of ‘data mining,’ wherein small subgroups of an unsuccessful trial are relentlessly scrutinized in search of groups for whom a benefit emerges, or seems to” [327].

The current US drug approval process does not ensure effectiveness, sometimes approving expensive “…tokens of healing as a cultural process” that are propelled by direct-to-consumer advertising until therapeutic failure is slowly demonstrated over years of use by the general population [328]. **Medical Ethicist Ezekiel J. Emanuel** noted that the FDA also approves drugs under an alternative, fast-track pathway that compares efficacy to no treatment (i.e., placebo or nothing), allowing “…interventions that are not ineffective, but offer minimal benefits for their cost” [329] (see **Appendix: 5.3 Pharmaceutical pricing and reform**).

“Harmonizing” dialysis

European firms and researchers appear to profit from the bad science (BS) in racialized US healthcare. European governments regulate drug prices and forbid misuse of ‘race’ at home [16], yet the US government lets European firms set prices and misuse ‘race’ to extract wealth that subsidizes, indirectly, European universal healthcare [330].

Two years after NASEM recommended against misuse of ‘race’ “…whether self-identified or not” [16], two groups of authors called for a new effort to “harmonize” ‘race’. In the *Journal of the American Heart Association*, they called “…to update recommended categories for global relevance” [331]. And in *JAMA Psychiatry*, they claimed “…that differences in reporting race and ethnicity across geographic locations… highlight the need for international guidelines to ensure equitable recruitment and reporting in clinical trials” [332]. Companies funding these authors include (among others) AstraZeneca, a Swedish-British company evaluating a drug for CKD aimed at *APOL1* RVs, and CSL Vifor Fresenius, a Swiss-German company providing 40% of US kidney dialysis treatments.

CSL Vifor Fresenius derives from North America <u>twice</u> the revenue per patient than from Europe, **Fig 18** [333]. The US government guarantees payment for kidney care [334]. Thus, neoliberal economics has US taxpayers supplying 80% of healthcare revenue to CSL Vifor Fresenius by paying double for the same service.

**Fig 18.**
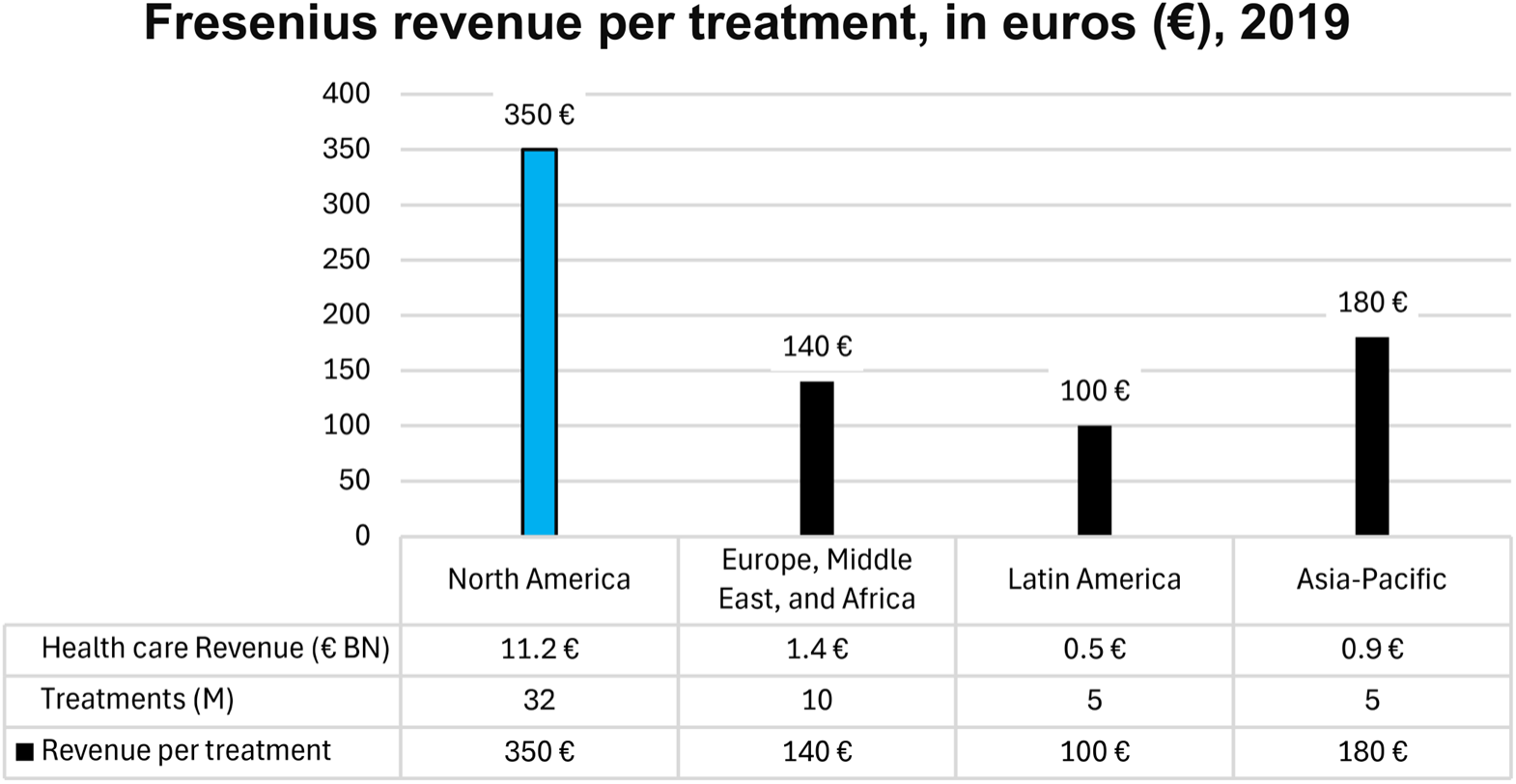
Fresenius reported marked differences in revenue per treatment across various regions (2019).

Diamonds and dialysis extract enormous wealth from the US, even though the former lack any intrinsic value [335,336]. Healthcare warrants a premium for superior outcomes, but US kidney outcomes are poor, **Fig 1**. The dialysis industry influences US policy to preserve profit [337,338,339], but the costly care is inferior:

> Taxpayers spend more than $20 billion a year to care for those on dialysis—about $77,000 per patient, more, by some accounts, than any other nation. Yet the United States continues to have one of the industrialized world’s highest mortality rates for dialysis care…. At clinics from coast to coast, patients commonly receive treatment in settings that are unsanitary and prone to perilous lapses in care…. Meanwhile, the two corporate chains that dominate the dialysis-care system are consistently profitable, together making about $2 billion in operating profits a year.
>
> One reason the system’s problems have evolved out of the health care spotlight is that kidney failure disproportionately afflicts minorities and the dispossessed. [340]

As **Editor-in-Chief of NEJM, Relman** included kidney dialysis in his 1980 warning against “…the medical-industrial complex” [341], while noting that health care is different from other services. However, no distinction was recognized when US policy shifted from trust in government to trust in for-profit businesses. In *The New York Review of Books*, Kuttner described the history:

> …a succession of Democratic presidents joined Republicans in turning away from the New Deal model of regulated capitalism toward what has come to be known as neoliberalism…. that markets work efficiently and that government attempts to constrain them via regulation and public spending invariably fail, backfire, or are corrupted by politics. As public policy, neoliberalism has relied on deregulation, privatization, weakened trade unions, less progressive taxation, and new trade rules to reduce the capacity of national governments to manage capitalism…. [342]

Neoliberalism grew healthcare to 18% of GDP by streamlining hospitals [343], opening land for reuse [344], providing ambulances [345], and exploiting unregulated US drug pricing [346] (see **Appendix: 5.3 Pharmaceutical pricing and reform**).

##### (d) Trust and institutional review

Finally, institutional review could invoke ethics to help end bad science (BS) and the misuse of medicontextual ‘race’ fueled by widespread conflicts of interest (COI) [347]. Until recently, research on trust in the medicontext of US healthcare primarily focused on “…trust issues related to systemic racism and health equity” [348]. A medical trope suggests that persuading Black patients to trust healthcare will reduce ‘racial’ disparities, but Warren et al acknowledged,

> …the deep and justified lack of trust that many Black Americans have for the health care system in general and clinical research in particular.… It would be wrong, as well as ineffective, to ask Black communities to simply be more trusting. [349]

Recent research described an inverse-u-shaped association between health literacy and trust—more trust at medium health literacy and less at low and high health literacy [350]. Often stereotyped as having low health literacy, Black Americans aware of their long history of abuse in US healthcare have one form of high health literacy, in a system that discourages expression of legitimate concerns [351]. **Ethicist Washington** noted, “…commentators have made Tuskegee an overburdened icon, suggesting that Black Americans are overreacting to a single study rather than to consistent abuse in the U.S. healthcare system” [352].

A more conventional form of high health literacy also supports ethical review, coming from those at the pinnacle of academic medicine.

###### Conflicts of interest

Three consecutive **Editors-in-Chief of The New England Journal of Medicine—Arnold S. Relman, Jerome P. Kassirer, and Marcia Angell**—linked doubts about medical research to COI. In 1980, **Relman** suggested that organized medicine should “…act decisively in separating physicians from the commercial exploitation of health care” to preserve its “moral authority with the public and the government” [341]. In 2002, **Relman and Angell** described the extent of pharmaceutical industry influence:

> It has also… assumed a role in directing medical treatment, clinical research, and physician education that is totally inappropriate for a profit-driven industry…. prescription drugs are not like ordinary goods, and the market for drugs is not like other markets. [353,354]

In 2015, a series of articles in *The New England Journal of Medicine* “…questioned whether the conflict of interest movement has gone too far in its campaign to stop the drug industry influencing the medical profession,” and **Steinbrook, Kassirer, and Angell** responded, “The problem, obviously, is that their objectivity might be compromised, either consciously or unconsciously, and there would be no easy way to know whether it had been” [355]. In 2016, **Angell** added, “Unless bias is blatant, there is simply no sure way to tell”, so we expect conflicted judges to recuse themselves, and we do not permit conflicted journalists to write the story: “Why should it be different for medical researchers?” [356].

The Strategic Council for Research Excellence, Integrity, and Trust of the National Academies expressed concern that: “Current processes for declaring an author’s relationships that might be viewed as potentially impacting his or her ability to address an issue without bias* are cumbersome, uneven, and inefficient” [357] (for example, printed journals that bury COI forms on a website), suggesting belief that one can be aware of one’s own bias, declare it, and then be truthful. Streamlining COI <u>disclosure</u> misses the mark in a world of monetized medical research and publishing. COI declarations shift the burden of discerning truth from publishers (with the leverage to ask questions and require compliance) to reporters and readers.

**Kassirer** noted, “…the vast attention paid to failure to disclose is misplaced… more attention must be focused on the financial conflicts themselves and their consequences” [358]. **Medical ethicists Joel Lexchin and Adriane Fugh-Berman** explained the real-world result:

> Disclosure of a conflict of interest, paradoxically, increases trust in the speaker, who is now viewed as an honest, forthright person…. Disclosure may lull audiences into believing, wrongly, that they can extract unbiased information from a biased presenter. [359]

Almost two-thirds of panelists developing clinical practice guidelines for kidney disease had financial COI [360]. Despite professional standards that editors, guest editors, and their staff should publish disclosure statements about their COIs, very few bothered to do so [361]. “Seventy-two senators and 302 members of the House of Representatives cashed a check from the pharmaceutical industry ahead of the 2020 election” [362].

When opioid deaths drew attention, congressional investigation found opioid money had compromised “…health professionals, patient advocacy groups, medical professional societies, research universities, teaching hospitals, public health agencies, policymakers, and legislators…”, creating the opioid crisis [363,364]. Annual opioid sales peaked at $8 billion (in 2015) [365]. Federal spending on kidney disease <u>increased</u> by $5 billion <u>annuall</u>y from 2015 to 2019, **Fig 2**.

When FDA approval of a drug rejected by outside experts drew attention, congressional investigation found FDA violated its own policies by collaborating on the company’s drug application and found internal company documents showing the company targeted physicians, patient advocacy groups, and other avenues to gain influence to have those groups apply pressure for FDA approval (as done by the opioid industry), while simultaneously seeking exemption from the Anti-Kickback Statute [366,367]. Unrelated to this drug, the company later offered $900 million to settle allegations of kickbacks to doctors in other areas [368,369]. Liu et al noted, “…pharmaceutical manufacturers penalized for kickbacks paid only 2.2% of US revenue accrued from selling implicated drugs during years of alleged violations…. a cost of doing business” [370]. (See FDA collaboration, **Appendix: 5.5.1 FSGS, COI, and BS**).

In 2026, a drug company offered academic researchers up to $125,000 for “…‘rigorous, independent’ work that focuses on the potential consequences of U.S. pricing policies on future innovation, the idea that pharmaceutical discovery is a strategic national asset…’ “ [371]. The pharmaceutical industry retained a former House Majority Leader to preserve unlimited prices for US taxpayers [372]. **ANGELL** argued otherwise [373] (see **Appendix: 5.3 Pharmaceutical pricing and reform**).

“The revolving door between the FDA and industry surprises few anymore” [374]. “Between 2013 and 2022, nearly two in five (38%) FDA advisory committee meetings included either a patient or consumer representative with active ties to a pharmaceutical-funded advocacy organization…. and in some cases, the organization received financial support from the product sponsor. These… organizational relationships extend beyond isolated cases and signal a broader problem warranting FDA policy attention” [375,376].

European industries extract wealth from ‘racialized’ US patients in ways their own governments forbid (e.g., misuse of ‘race’ in drug approvals [7,322]) or regulate (e.g., drug prices) to provide universal healthcare that the US does not [18]. An article promoted eGFR for early detection of kidney disease [377] despite its unacceptably high eGFR false positive rate above 60 mL/min that favors diagnosis using low-cost, widely available creatinine [1,378]. Its authors declared “…no conflict of interest to report in relation to the topic of this study”, but several indicated funding from, for example, AstraZeneca and Fresenius (part of CSL Vifor), two European companies that would be affected by reliable early diagnosis of CKD to prevent progression to KF.

International reviewers may understand, consciously or unconsciously, that they benefit from US structural racism. A European reviewer funded by CSL Vifor Fresenius withdrew support for our preCKD manuscript after revisions addressing his comments explored ethics of ‘race’ and COI (see Comments to Version 9 of our preCKD manuscript [1]).

This is not to name a handful of bad actors but an example of an underlying pattern of bad science (BS). There is financial incentive for healthcare businesses to help researchers continue work that enables their business interests, so they redirect some of that wealth. Even without owning shares in a healthcare corporation (i.e., no direct gain from rising stock price), researchers with no current relationship to industry may hope that future funding will keep them employed. Such influence is not captured on a COI form.

###### Good funders make good science

Governments may fund research into low-cost options, but a venture-capitalist physician advised rejecting <u>anythin</u>g without market potential. For example, low-cost, widely available serum creatinine can detect early CKD [1], but eGFR cannot [1]. Ignoring creatinine allows eGFR to preserve the preCKD blind spot for eventual replacement, in the lucrative US market, by a new test that will be unaffordable in large parts of the world.

In *Reason To Believe: How and why irrationality takes hold, and what do to about it*, **Steven Pinker, Professor of Psychology at Harvard University,** described the rationale for disinterested government funding of science:

> How, then, can we explain the pandemic of poppycock? My best answer comes in four parts.
>
> The first is rooted in the very nature of rationality. Reason, almost by definition, is inference deployed in service of a goal; no one gets rationality credit merely for enumerating true but useless propositions. But that goal need not be an objective understanding of the world. It can also be to win an argument in which the stakes matter to you. As Upton Sinclair pointed out, “It is difficult to get a man to understand something when his salary depends upon his not understanding it.”
>
> Or reason can be used to show how wise and moral your group is—your religion, tribe, your political sect—and how stupid and evil a rival one is. This “myside bias,” explained in Keith Stanovich’s *The Bias That Divides Us,* appears to be the most pervasive bias among the hundreds documented by cognitive science and social psychology. And there’s a perverse rationality behind it. It’s far from irrational for an individual to endorse a belief that gains them status as a hero for their side while avoiding ostracism as a traitor. The problem is that it’s irrational for a society as a whole to lurch between one faction’s dogmas and another’s rather than collectively arrive at the most accurate understanding of reality. These conflicting incentives place us in a tragedy of the rationality commons. [379]

Funders with an agenda generate bad science (BS). In *The New York Review of Books*, **Angell** described the effects of industry funding on peer-reviewed research:

> Because drug companies insist as a condition of providing funding that they be intimately involved in all aspects of the research they sponsor, they can easily introduce bias in order to make their drugs look better and safer than they are…. In short, it is often possible to make clinical trials come out pretty much any way you want, which is why it’s so important that investigators be truly disinterested in the outcome of their work….
>
> Conflicts of interest affect more than research. They also directly shape the way medicine is practiced, through their influence on practice guidelines issued by professional and governmental bodies, and through their effects on FDA decisions….
>
> In recent years, drug companies have perfected a new and highly effective method to expand their markets. Instead of promoting drugs to treat diseases, they have begun to promote diseases to fit their drugs. The strategy is to convince as many people as possible (along with their doctors, of course) that they have medical conditions that require long-term drug treatment. Sometimes called “disease-mongering,” this is a focus of two new books: Melody Petersen’s *Our Daily Meds: How the Pharmaceutical Companies Transformed Themselves into Slick Marketing Machines and Hooked the Nation on Prescription Drugs* and Christopher Lane’s *Shyness: How Normal Behavior Became a Sickness*.
>
> To promote new or exaggerated conditions, companies give them serious-sounding names [e.g., “brain fog”, “the kidney-failure gene”]….
>
> It is simply no longer possible to believe much of the clinical research that is published, or to rely on the judgment of trusted physicians or authoritative medical guidelines. I take no pleasure in this conclusion, which I reached slowly and reluctantly over my two decades as an editor of *The New England Journal of Medicine*. [380,381]

Widespread COI and the replication crisis signaled that “Science is in trouble” [382]. **Richard Horton, Editor-in-Chief of The Lancet** [383] summarized,

> The case against science is straightforward: much of the scientific literature, perhaps half, may simply be untrue. Afflicted by studies with small sample sizes, tiny effects, invalid exploratory analyses, and flagrant conflicts of interest, together with an obsession for pursuing fashionable trends of dubious importance, science has taken a turn towards darkness. [384]

In 2026, **Ioannidis** described the challenge,

> More than 50% of online content is now written or translated by AI, not humans. Scientific articles, for example, can now be written within seconds. Perhaps the quality of these fabricated materials is justifiably questioned, but by comparison, the “genuine” human-generated bibliography is not a heap of diamonds. It looks more like a garbage dump where here and there some remarkable achievements are found among the debris. The annual number of fake articles produced by AI may now exceed a million annually (one in 7 submissions). The number of trivial articles that we don’t even need to ask ourselves if they are wrong or fake (since they are simply useless) is probably many millions. [385]

In 2014, **H. Robert Horvitz, Professor of Biology at MIT and Nobel Laureate in Physiology or Medicine**, addressed a joint session of 400 students of science from three urban public high schools [386] (a student remarked, “No scientist <u>ever</u> came to talk to us before”). Describing for the students not-yet-published research on *C. elegans* “tasting light” [387], **Horvitz** noted the crucial role of government in funding good science:

> ‘Basic research,’ Horvitz said, ‘is the driver of scientific knowledge.’ Private companies have little incentive to pursue basic research, so the government is the primary source of its financial support…. It is important that governments understand how research contributes to the function of everyday life, he said. [386]

In 2025, **Angell, Kassirer, and Steinbrook** noted a key element to restore trust in science:

> …researchers must wean themselves almost entirely from drug company money. And they can do this only if they have a reliable alternative source of funding. If we want U.S. scientists to rely less on pharmaceutical dollars, we must make more NIH or other governmental dollars available. [388]

The task is dauntingly impossible, but for clinicians who no longer know what to believe, **Ethicist Washington** suggested resources:

> Several books offer clear, impeccably researched guides to sniffing out manipulation. Angell’s *The Truth About the Drug Companies* and Abramson’s *Overdosed America* are likely to be most helpful to a busy clinician. The PharmedOut website offers tools for detecting undue influence in medical research and publishing. [327]

###### Institutional review

It is a fair question whether and when collecting ‘race’ is worth the risks [389,390], and whether those risks can be avoided by introducing ethical regulatory oversight. **Medical ethicists Raymond G. De Vries and Scott Y. H. Kim** offered hope at the intersection of sociology and bioethics:

> The collaborative work that gets done under the rubric of empirical bioethics moves the important ideas of philosophical bioethics into the real world of medicine and medical research where human beings live and work, and help and harm each other. [391]

Institutional review boards (IRBs) could challenge misuse of ‘race’ under existing regulations that encourage protection of those “…vulnerable to coercion or undue influence” [392,393,394,395]. However, recent developments risk weakening ethical oversight. In 2017, NIH adopted a single IRB (sIRB) model and eliminated local IRBs from multi-center trials. **MEDICAL ETHICISTS ANN-MARGRET ERVIN, HOLLY A. TAYLOR, AND STEPHAN EHRHARDT** cautioned, “We do not yet know whether streamlining IRB review will affect an IRB’s ability to protect the welfare of human participants” [396] (see **Appendix: 5.4 ‘Race’, risk, and novel research**). Although commercial and non-profit IRBs are <u>both</u> susceptible to COI (and how to regulate them is debatable [397]), **medical ethicist Arthur L. Caplan** noted,

> ‘This shift to commercial IRBs is, in effect, over,’ said Caplan, who heads the division of bioethics at New York University Langone Medical Center…. Commercial IRBs now oversee an estimated 70 percent of US clinical trials…. with larger IRBs buying smaller ones, and even private equity firms coming along and buying the companies…. which often have harder-to-spot conflicts of interest and a profit motive. [398]

###### Summary

We have provided a discussion of bad science (BS) to show that equal outcomes should be expected under equal conditions regardless of ‘race’. We reviewed how ‘race’ is misused, manipulated, and selectively included or omitted. We described international ‘racial’ inconsistencies, which are internalized from childhood experiences, and showed that medical researchers consciously and unconsciously spread the myth of ‘race’. We noted that Black people rationally distrust medicine and medical research. Given FDA regulations promoting ‘race’ and the power asymmetry between subject communities and the pharmaceutical industry, it seems unlikely that collaboration alone [399] will temper the profit motive in misuse of ‘race’.

American bioethics favors autonomy almost to the exclusion of its other three founding principles, but as **medical ethicist Joseph J. Fins** noted, we should “…move beyond narrow questions of patient choice, particularly when the disenfranchised are not in a position to exercise that choice” [400]. Fins added, “…that while rights are necessary to avoid exploitation, they are not sufficient and that instead there should be a focus on capabilities that promote human flourishing” [personal comm]. In this case, it is about identifying nascent kidney disease with metrics that are accurate and accountable to scientific and ethical norms [1,16]. Waning professional independence is no match for the incentives and economic power in unregulated profiteering. Further research might show whether medicontextualism and an appeal to international bioethics could end bad science (BS) and the misuse of ‘race’ in medical research.

## 4. CONCLUSION

Medicontextualism in re-analysis of ‘race’ reveals bad science (BS) largely stemming from cross-cultural conflicts in its social medicontexts such that research associating socially constructed ‘race’ with any explanation other than structural racism will always be wrong. Simpson’s paradox prevails when combining veterans with the general public (or Philadelphia, PA, with Charlotte, NC, **Fig 4**) in the ‘racial’ bad science of “harmonization” and homogenization.

Selection and training of servicemembers create comparable starting conditions for veterans. Those in Veterans Administration Healthcare showed comparable social determinants of health and comparable kidney outcomes, which should be the expected norm for all under more equal social conditions—regardless of socially constructed ‘race’, ethnicity, or nationality. Further research to identify essential veteran cofactors may improve general population health, reduce ‘racial’ disparities, and redirect some resources to prevention.

Genomic studies searching for biological ‘racial’ differences overlook pigmentary genes at the root of medicontextual racism and its socially constructed environmental, educational, and economic gradients. Apolipoprotein L1 “high-risk” variants are the latest proposed “cause” of the “genetic” form of focal segmental glomerulosclerosis. However, the increased risk appears too small to justify treatment at the population level, and *APOL1* (the target gene-of-the-day) may be an innocent-neighbor proxy for a pigmentary gene. Broad application of nonspecific, medicontextual Population Health measures may further mitigate risk.

Conflicts of interest, single IRBs, and risks in novel research warrant review of the assumptions in ethical oversight.

Despite scholarly consensus and 100 years of academic articles that biological ‘race’ is bad science (BS), misuse persists in medical research, suggesting, for further study, we may need to **(1)** work together, **(2)** discourage standing in the doorway, and **(3)** appeal to principles of international bioethics to end misuse of ‘race’.

## 5. APPENDIX

### 5.1 Ethics statement

**‘Race’ in research:** NASEM advised, “Researchers should be as transparent as possible about the specific procedures used to name groups within their data sets” [16]. We did not present ‘race’ as an input to clinical decisions, a means of stratifying care, or an hypothesis for genetic difference without data. One primary source used ‘race’ designated in Medicare Denominator File or VA data sources [73]. We explored the approach to ‘race’ and ancestry in the other primary sources (see **Discussion**).

**Human subjects:** The primary sources used information collected for non-research activities in VA Healthcare or obtained informed consent from all participants after receiving IRB approval. Secondary re-analysis of publicly available data is not considered human subjects research and is exempt from review [401].

**Privacy:** The study included no individually identifiable patient health information.

### 5.2 Methods

Our main hypothesis is that because ‘race’ is socially constructed, social context and social cofactors will remove or explain ‘racial’ differences in CKD and KF. A corollary is that medical researchers trapped in a world of ‘racial’ illusion create flawed research, then publish under a system that perpetuates the myth of <u>biologic</u> ‘race’ by actively blocking examination of medicontextual root causes. Therefore, re-analyzing studies that endorse <u>biological</u> ‘race’ will <u>always</u> be fruitful. During recent questioning of eGFR ‘race corrections’, our preliminary re-analysis of a study of KF in Black and White veterans (**Section 2.1**) elicited responses referencing *APOL1* (e.g., “the kidney failure gene”) as “validating” ‘racial’ difference in kidney disease. We broadened our literature search.

Relevant to our hypothesis, we searched PubMed (on February 24, 2024) for titles or abstracts that included “veteran” and “APOL1”, yielding four studies [402,74,78,279]. Of three studies directly addressing kidney disease, we added re-analysis of two (**Sections 2.2 and 2.3**) and noted the third in **Section 3.3.2 Racism’s genome-wide associations**. We selected these articles because they represented an emerging program of research on the genetic underpinnings of ‘race’ and kidney disease. While it may not represent all studies on this topic, the goal was to illustratively characterize how the body of literature may support the medicontextual myth of ‘race’ as biology.

Our secondary re-analyses involved extracting primary data of veterans from previously published tables and figures (i.e., data of all subjects in two of the studies and 80% of subjects in the third), using knowledge of socially constructed ‘race’ and obvious medicontextual social cofactors to reorganize the data into simple, accessible graphs and charts to better compare RRs and ORs of CKD rates and KF outcomes.

#### 5.2.1 Re: Removing ‘race correction’

The large study of KF in veterans by Choi et al [73] presented enough data to reverse the eGFR ‘race correction’. We repurposed data from its first and second tables, removed ‘race correction’ by dividing MDRD eGFRs by 1.21, and examined eGFR prevalence and rates of KF against eGFR results with and without ‘race correction’. Mathematically transforming a variable by a constant has a one-to-one impact on the magnitude of the regression coefficient [403], and the estimated association will change proportional to that number, which may be useful if an actual difference could be overlooked. However, if no true ‘racial’ difference exists, the association between eGFR prevalence rates and KF rates for White and Black patients should be similar, and ‘race correction’ would inflate the association’s magnitude, implying more KF than found.

We estimated the association among White patients, Black patients with unadjusted eGFR rates, and the same Black patients with eGFR ‘race correction’ removed, hypothesizing that the ratio of associations for ‘race-corrected’ data and ‘not-race-corrected’ data would equal 1.21, the exact magnitude of ‘race correction’. Additionally, we hypothesized that there would not be a significant difference in associations between White and Black patients after removing ‘race correction’.

#### 5.2.2 Re: Veterans and APOL1

**Re: *APOL1*, AKI, and COVID:** We re-analyzed a study by Hung et al [74], calculating RR and OR from primary data in their first and second tables. Although we dismissed this article in an early version of this manuscript (due to uneven distribution of vasopressors and mechanical ventilation between those with and without AKI), repeated encounters with references to it as “proof” that genetic susceptibility causes medicontextual ‘racial’ disparity prompted our full re-analysis and PubMed search.

We adjusted the *APOL1* RR to remove the confounding of high-impact cofactors. For comparison, we included a similar study in the Brazilian general population [77], where ‘race’ is defined differently. Because some CIs were unusually asymmetric, we recalculated them from the given p values and z-statistics.

**Re: *APOL1* and N264K:** We similarly re-analyzed another study by Hung et al [78] that examined associations between *APOL1* LR and HR genotypes, N264K variants, and the US taxonomy of socially constructed ‘race’. We focused on the data from US veterans (the bulk of their data) and created simple graphs to better show the relationships within the data. Because N264K**+** is an *APOL1* variant more common in Europe (occurring in less than 5% of US Black veterans), we further narrowed to the 95% of Black veterans without N264K, calculating and ordering results by ORs between the LR and HR (N264K**-**) subgroups.

### 5.3 Pharmaceutical pricing and reform

“The U.S is unusual in that it lacks any centralized mechanism for negotiations between payers and drug manufacturers” [404]. Most countries regulate the price of medicines, but in the US, international drug companies charge whatever Americans will pay [405,406]. While regulating them at home, European governments did not object to harmonized ‘race’ and extreme pricing in the US, preserving the US drug market as the most lucrative in the world. In 2018, **ETHICIST EMANUEL** noted, “The whole world spends about $1 trillion on drugs, and roughly half of that is spent in the U.S.” [407]. In 2022, as 4.3% of global population, we paid 62% of brand name drug costs, **Fig 19** [408,409].

**Fig 19.**
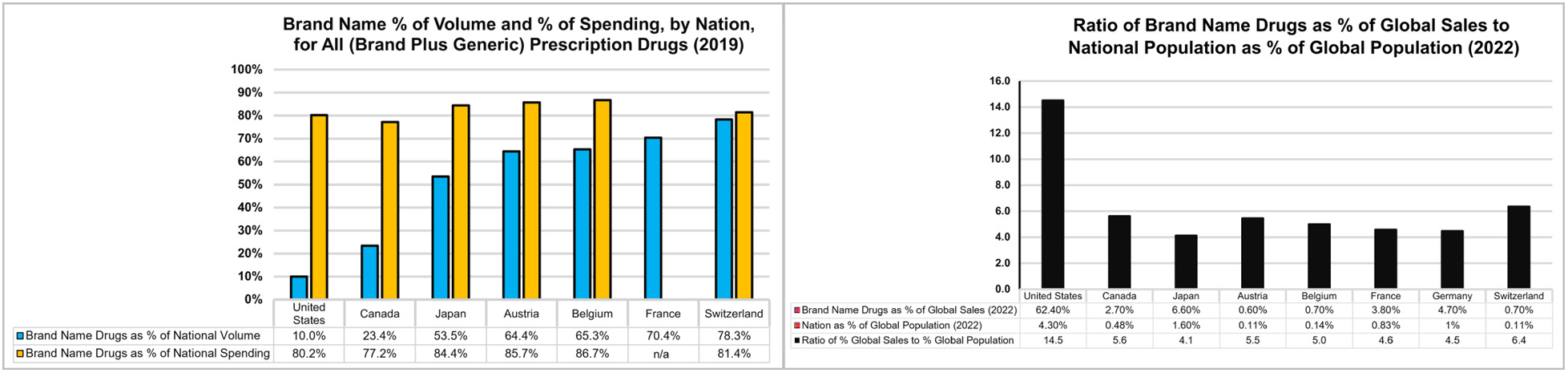
Visualizing the extreme expense of brand name drugs in the US (left): The US is one of the few high-income nations that do not regulate drug prices. Brand name drugs are only 10% of US drug volume but absorb 80% of US drug expenditures, **(right):** The ratio of US brand name drugs (as a % of global drug sales) to US population (as a % of global population) is far higher than for other industrial countries, where drug prices are regulated, yet pharmaceutical companies are still profitable.

For example, FDA approved semaglutide (Ozempic, Wegovy) for type-2 diabetes, then weight loss. The estimated production cost was $193 to $380 per person-year, a small fraction of branded semaglutide’s price: “…$6,936-7,716 per person-year… prior to recent price increases of… 3.5% in 2025” [410].

In 2002, **Relman and Angell** noted,

> The ten American pharmaceutical companies in the Fortune 500 list last year ranked far above all other American industries in average net return, whether as a percentage of revenues (18.5 percent), of assets (16.3 percent), or of shareholders’ equity (33.2 percent). (For comparison, the median net return for other industries was only 3.3 percent of revenues.) [353]

A series of regulatory changes [411] led the pharmaceutical industry to raise prices by referencing the estimated value of a quality-adjusted life year (QALY) [412,413,414]. Value-based drug pricing takes responsibility for bundled services. A drug does not save a life by itself but requires support services. These services include drug delivery systems, medical and nursing staff, health facilities, primary care, food, housing, and “…public health and social measures” [415], but in contrast to extraction of profits to distant stockholders, most payments for support services circulate several times in the local economy as wages are spent. If not “bundled pricing” for healthcare efficiency [416], then other unregulated vendors and services could extract as much (or more) for their contributions to the same life. If bundled pricing, then not reimbursing for the support services is “unbundling” [417].

In her closing chapter, *How to Save the Pharmaceutical Industry—And Get Our Money’s Worth*, **Angell** wrote:

> Despite all its excesses, this is an industry that should be saved—mainly from itself. The public is dependent on it, and it should be made to carry out its original purpose of developing important drugs and selling them at reasonable prices. I have shown how the industry, corrupted by easy profits and greed, has deceived and exploited the American people. But that is not to say that everyone who works for big pharma is corrupt or deceitful. In fact, it is my impression that most pharmaceutical employees, even at the highest levels, accept their own public relations. They honestly believe they are part of an innovative industry whose prices are an accurate reflection of the value of its products and the costs of making them. That is a testament to the effects of compartmentalization in big corporations; very few people know the full dimensions of the business. And it is also a testament to human nature. People want to be proud of their work.
>
> But readers will know by now that the pharmaceutical industry, despite its many dedicated employees, has moved a very long way from its original mission. In this chapter I will propose specific reforms that would restore the industry to its purpose and make prescription drugs not only more affordable but better and safer. My intention is to show what needs to change to make the industry function as it claims to—as a source of innovation and affordable drugs to help people lead longer, better lives…. [373]

### 5.4 ‘Race’, risk, and novel research

The US National research Act of 1974 initiated research governance and led to the “Common Rule”, which sometimes protects patients from becoming unwitting subjects in research: “…There are many research activities that do not come under the Common Rule. An example is research funded by private money….” [418].

New research methods expose <u>all</u> patients to ethical lapses, including Black patients, who are overrepresented in dialysis centers and emergency rooms—the last resort for uninsured people. In this section, we discuss the ethical limitations of including ‘race’ in the process of doing research.

#### 5.4.1 Eroding informed consent

**Ethicist Washington** described gradual erosion of the first principle of the Nuremberg Code, informed consent:

> …researchers bypassed eliciting subjects’ consent by obtaining waivers of informed consent for studies on a case-by-case basis. Then a 1996 law… provided a waiver for certain types of medical research said to carry only “minimal risk.” …
>
> …another 1996 change in the Federal Code of Regulations: withholding informed consent from unconscious trauma victims [419,420,421,422]….
>
> This approach is an ethical error. If a person (or her legally appointed representative) cannot give consent, she should be treated with duly tested standard care as if there were no experiment to worry about. [352]

Pragmatic clinical trials (PCTs) or cluster randomized trials (i.e., on clusters of populations served by distinct clinics or hospitals) raise novel ethical issues [423]. The fact of a study may be obscured to the subjects by blanket waiver of informed consent when the IRB finds “minimal risk” and that the waiver does not “…adversely affect the rights or welfare of the participant” [424]. Disagreement between “risk-averse” and “permissive” IRBs on multi-site studies is avoided by allowing only one review, by a possibly remote sIRB, and approval may seem justified if the subjects experience “…a minor increase over minimal risk” [425]. However, because the determination of “minimal risk” by either type of IRB might be incomplete, another approach is to require dialogue among IRBs representing their local populations, aiming to understand and resolve any fact-dependent disagreements among their experts [426].

Some ethicists questioned the lack of public oversight of such studies [427] and the creeping application of waiver of consent to studies manipulated to fit into waiver criteria [428]. These practices undermine autonomy of patients and clinicians alike. In a study of attitudes toward PCTs of maintenance dialysis in patients with kidney failure, subjects and their partners preferred to be given the choice before engaging in such studies, even anticipating loss of trust if the study was not openly declared [429]. That a “substantial” minority of patients might accept a hypothetical study done without written consent [430] suggests confounding by “therapeutic illusion” (belief that a study is done to benefit the subjects rather than future populations). Common law and state statutes require informed consent, even for “standard of care” medical treatment [431]. And as **Medical ethicists Shona Kalkman, Ghislaine J. M. W. van Thiel,** et al noted, even “stakeholders” saw

> …ethical challenges in four domains: (1) less controlled conditions creating safety concerns, (2) comparison with suboptimal usual care compromising clinical equipoise, (3) tailored or waivers of informed consent infringing patient autonomy, and (4) minimal interference with real-world practice reducing the knowledge value of the results”. [432]

#### 5.4.2 Declining physician autonomy and community influence

Clinician autonomy affects trustworthiness [433,434]. Most clinicians have now ceded their autonomy to corporate interests [435,436] and the well-intentioned corporate impulse to stifle dissent [437]. Clinicians are expected to see more patients in less time, at lower cost, with better outcomes, under greater oversight, and at higher patient satisfaction [438,439], while inundated with medical literature they cannot trust and admonished to “Get With The Guidelines” [440].

No longer admitting their patients, primary clinicians lost eligibility for the hospital medical staff, ending inpatient care by those who could best advocate for their patients. Pushed to be “efficient”, hospitals hired clinicians dedicated to inpatient care, with incentives to shorten length of stay. Hospitalists now supervise young, highly educated doctors-in-training, who decide when a patient is “not too sick to go home”, or when care at 75 might be “wasting resources”, or when a family should be encouraged to “request” opiates for “comfort” because “…it would be inhumane to continue”. Actions ethically questionable under Saint Thomas [441] are now okay in the name of private equity. Shorter length of stay suggests savings but may move costs from one bottom line to another: ambulance ride, daily bed fees at the next facility, and hidden costs and risks in mid-treatment transfer of care to new medical and nursing teams. Demanding homecare from families with uneven resources may subtly contribute to ‘racially’ disparate outcomes. Not long ago, highly valued “continuity of care” had primary physicians making morning and evening hospital rounds to supervise inpatient care. Familiar with patient and family, they could sometimes keep a patient in hospital till well.

Clinicians dedicated primarily to patient care, “…whose primary interest is your… welfare (rather than following a research protocol)”, are different from medical researchers [431]. The declining influence of community physicians and loss of independent voices on the medical staff eroded an informal layer of ethical review, reducing communication between hospital and community that is a crucial consideration for PCTs [442], particularly when a possibly remote sIRB might declare “minimal risk” or “a minor increase over minimal risk” that allows blanket waiver of informed consent.

#### 5.4.3 Shifting IRB review

Ethicists can disagree, so seemingly redundant review by IRBs at several sites can improve understanding of ethical risks, like “minimal risk” [423,424,425,426]. Nevertheless, without legislative input by the US Congress or executive input by the Office of Civil Rights, government scientists loosened their own oversight rules and mandated a sIRB for multi-site human subjects research, which favors commercial IRBs. **Medical ethicist George J. Annas** noted the significance of IRB review:

> Although IRB review was not required by the Nuremberg Code… doing away with the requirement for all but 1 institution involved in a multicenter research trial could gut the review requirement of any real meaning…. because the structure of the regulations requires holding the individual institution “responsible for safeguarding the rights and welfare of human subjects.” … The single IRB requirement risks replacing ethics with efficiency….
>
> Under existing regulations, all members of an IRB but 1 could have an inherent conflict of interest because they can be employed by or affiliated with the institution. [443]

In public comments on the NIH sIRB policy, medical researchers welcomed research efficiency [444]. We condensed some ethical concerns into **Fig 20a** and **Fig 20b**. Reduced oversight contradicts the original impetus for research governance—loss of faith in medical altruism:

**Fig 20a.**
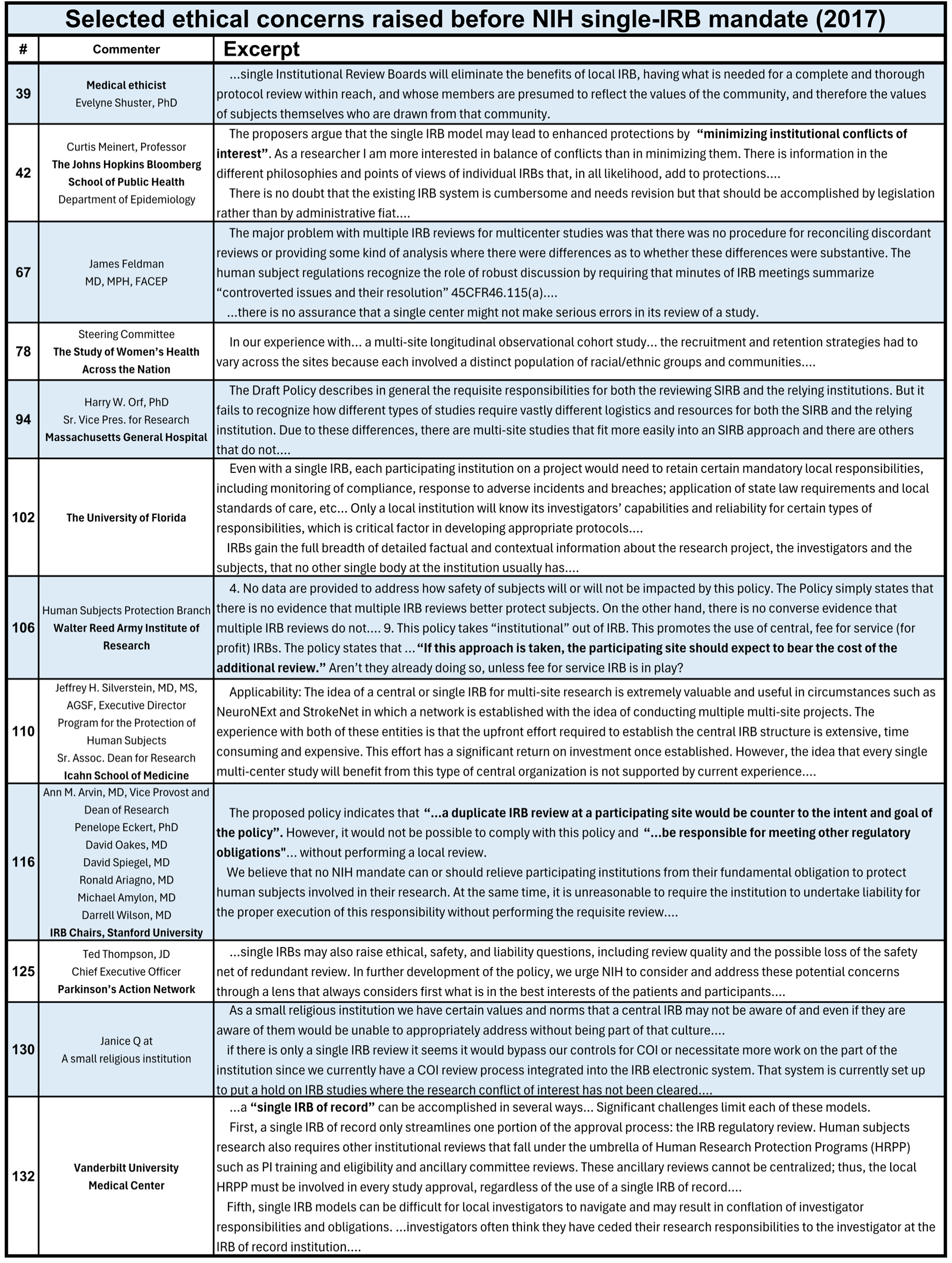
Selected ethical concerns raised before the NIH single-IRB mandate (2017).

**Fig 20b.**
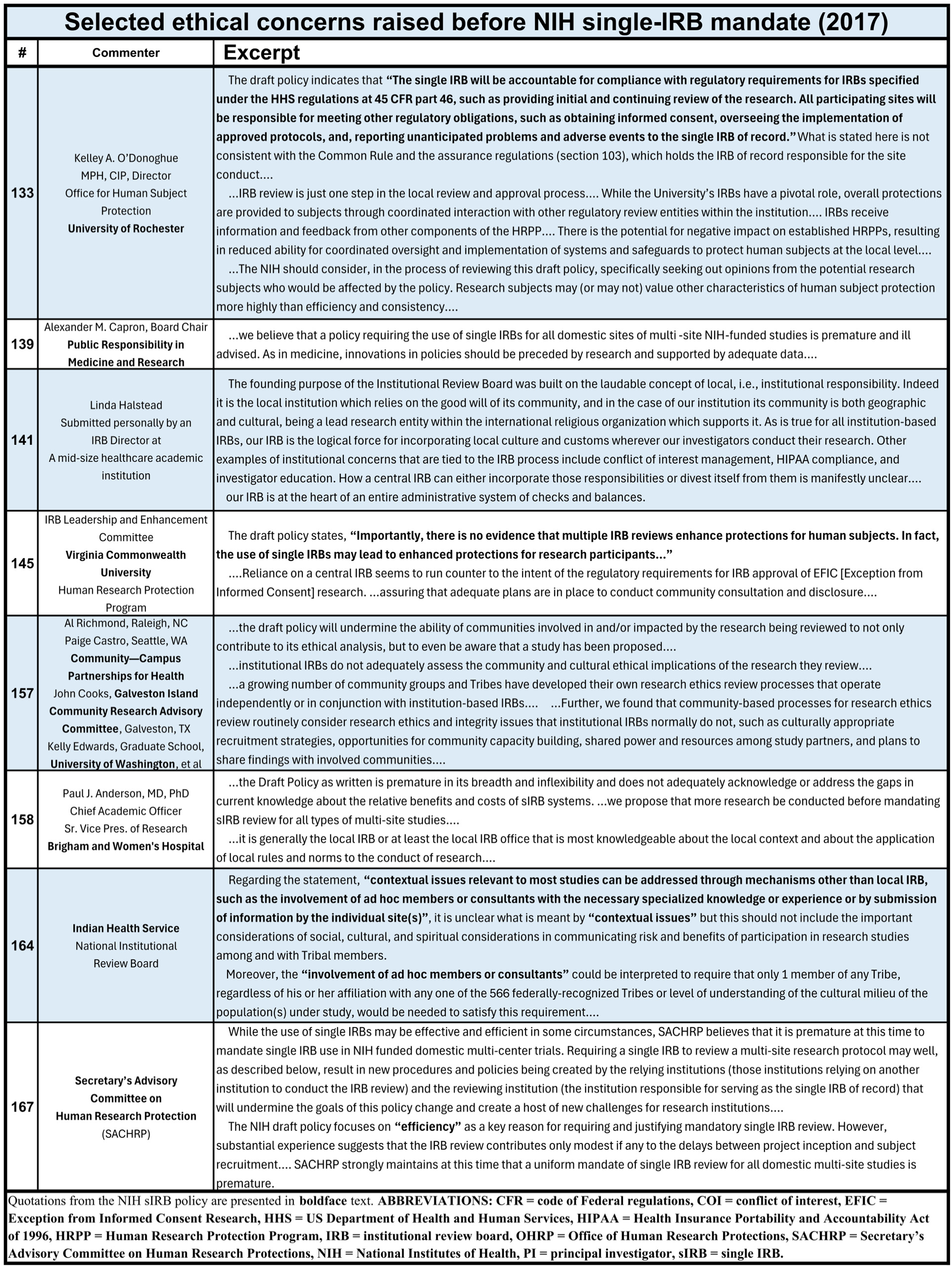
Selected ethical concerns raised before the NIH single-IRB mandate (2017).

> …the altruistic nature of medical care was woefully insufficient protection for patients even after such horrific revelations as the Nuremberg trials, the Tuskegee syphilis study, Dr. Beecher’s exposé [445], and other such reports. These revelations engendered such a distrust of altruism in medicine, that federal legislation…. codified protection of human subjects requiring a structure and process for institutional review boards to approve all human subject research. [446]

Oversight of that structure and process for institutional review boards continues to change. The Secretary’s Advisory Committee on Human Research Protections “…that advises the Department of Health and Human Services on emerging ethical and legal issues in human health research has been disbanded” [447] (see **Fig 20b**, comment # 167), and federal oversight by the Office of Human Research Protections has been limited, chronically, “…by anemic budgets and staffing challenges that complicated its mission to ensure compliance with federal ethics and safety regulations across multiple administrations” [448]. The Common Rule—the federal regulations for the protection of human research subjects—applies only to studies that accept federal money. Further, **MEDICAL ETHICISTS KYLE B. BROTHERS, SUZANNE M. RIVERA, JEAN CADIGAN, RICHARD R. Sharp, and Aaron J. Goldenberg** noted,

> …existing frameworks for regulation and oversight of research with human participants fit poorly with contemporary research…. [T]he 2011 to 2017 process undertaken to ‘modernize’ the Common Rule…. did not adequately consider the transformation that has occurred with regard to cultural perspectives on a range of critical issues relevant to the oversight of research with human subjects. These include, but are not limited to: changing perspectives on the right to privacy (and limits thereto), power imbalances between experts and lay people, trust in institutions, and individual ownership and control of data and biomaterials. [449]

**Medical ethicists Stephen Rosenfeld and Holly Fernandez Lynch** advocated reforms to reduce COI [450] and cautioned,

> IRB determinations are only one step in a very complex system to protect human subjects and the step possibly furthest removed from the actual, day-to-day conduct of research where harms are realized. The mistaken view that IRBs are the primary mechanism to protect research participants against harms has consequences for the debate about IRB quality. In particular, such a view suggests that IRBs should be assessed against the impact of their review on reducing actual harms to research participants rather than against the completeness and integrity of the review itself. In fact, IRBs are charged with determining that “Risks to subjects are minimized” and that “Risks to subjects are reasonable in relation to anticipated benefits.” [451,452]

Ethicists can disagree, so one feature of past practice—redundant IRB review at multiple sites—appears especially important in the subjective assessment of “minimized” or “reasonable” risks. Ongoing misuse of ‘race’ raises the risk and diminishes the “anticipated benefits” for Black patients, as noted by **ethicist Washington** [302]. Thorough review by more people from diverse locations and backgrounds broadens the values weighed and increases the chance for alternative experiences to be heard and considered. Our research suggests risks that may be unknown or ignored by IRB members, a prospect more likely under the sIRB model. If the belief that IRBs exist to protect patients is mistaken, then it may be time to revise the National Research Act.

Whether IRB review provides ethically sufficient protection for patients unaware of medical research when regulations waive written consent is unclear. Waivers are made for practicality—it is difficult to obtain consent and enroll large numbers of participants in research studies, so it is easier to streamline the process. However, because non-researchers often lack detailed understanding of what research is, how research data are used, or representation on IRBs, it begs the question of whether this streamlining represents the best interests of the community being studied or just makes it easier for researchers to engage in research.

### 5.5 Medicontextual FSGS and APOL1

Bad science (BS) promotes arbitrary distinctions that may seem unrelated to medicontextual exposures of structural racism. FSGS and APOL1 are two examples of invalid conclusions presenting Black patients as outliers.

#### 5.5.1 FSGS, COI, and BS

In May 2023, a drug targeting FSGS failed its two-year clinical trial: “…there were no significant between-group differences in eGFR slope at 108 weeks” [453]. Yet, in April 2026, the FDA approved Filspari for a subgroup of “…patients with FSGS and without nephrotic syndrome”. <u>After</u> the clinical trial results, a professional society funded by the Filspari manufacturer (Travere Therapeutics) had advocated “…the use of proteinuria reduction measured over two years as a trial endpoint that regulatory agencies should use as the basis for approving drugs to treat FSGS….” [454]. The drug’s manufacturer funded a patient advocacy group (as done by the opioid industry) that also advocated for the drug. Finally, the FDA helped the company, by suggesting subgroup analysis to meet the post hoc standard, then approved the drug, turning scientific failure into marketing opportunity as “the first drug to treat FSGS” [454]. Subgroup analysis is rarely confirmed in follow-up studies [455,456,457,458,459].

##### Primary nephrotic syndrome

Although it did not slow CKD progression and was not approved for FSGS in the setting of nephrotic syndrome, Filspari will likely be promoted in a way that leads to prescribing outside its approved use—even greater risk of ‘racialized’ use.

First, **HOWIE** showed the confusion in FSGS histology [221]. We discussed FSGS and APOL1 in **Section 3.2.6 Focal segmental glomerulosclerosis**.

Second, a retrospective review of adult primary nephrotic syndrome over 10 years, at a hospital system in Metropolitan New York, tallied the causes of primary nephrotic syndrome by ‘race’ and reported, “…a diminished frequency of focal segmental glomerulosclerosis in non-black patients” (i.e., Black patients were different) [460]. In the four columns to the left of **Fig 21**, the original data shows Black patients as outliers.

**Fig 21.**
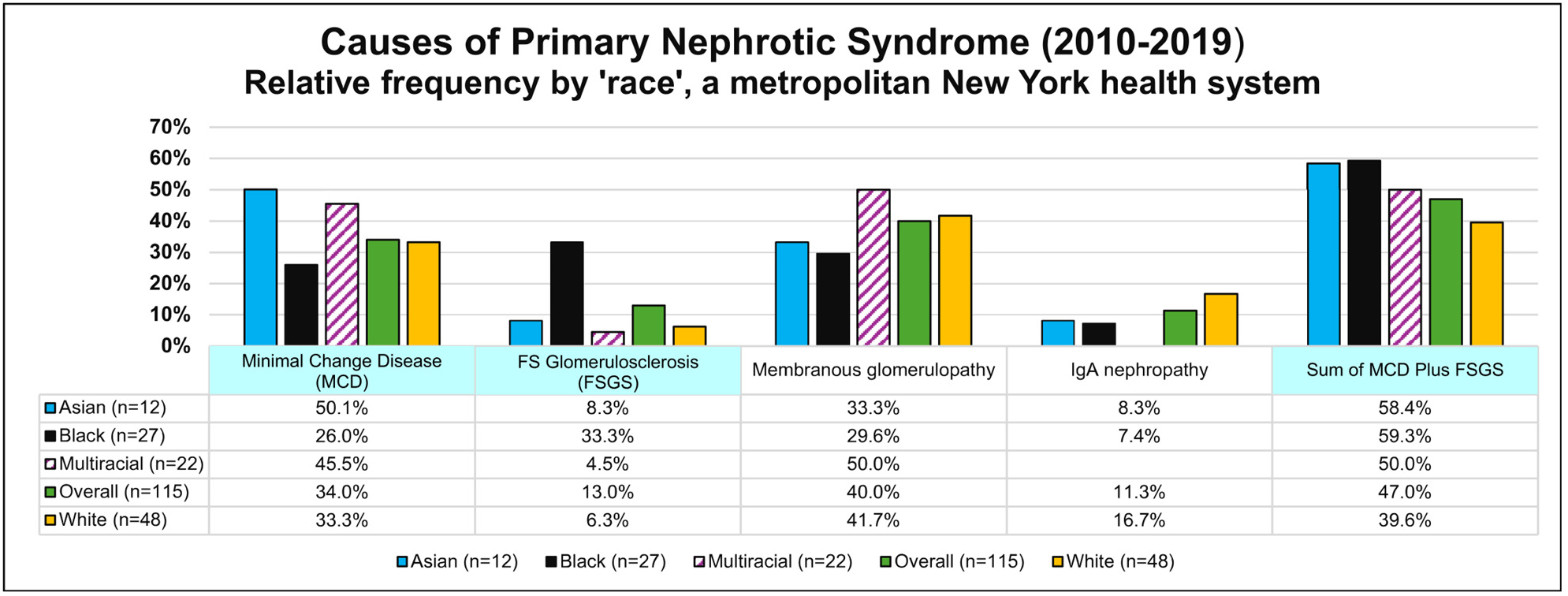
Causes of primary nephrotic syndrome by ‘race’. Note that although Black patients appear different when causes are separated into subgroups defined by severity, summing the histopathological subtypes of minimal change disease (MCD) and focal segmental glomerulosclerosis (FSGS) reveals a similar distribution for all patients (three columns to right). Differences in the two left columns may reflect later presentation, after less early care (under “race corrected” eGFRs and past belief that Black patients had less early kidney disease, so needed less early nephrology care), leading to the belief (and reality) that, far more often than other ‘races’, Black patients were diagnosed and treated late, presenting with kidney disease that often progressed rapidly to kidney failure. The concept of “*APOL1* FSGS” blames this ‘racial’ disparity on genetics, obviating need to search for environment and SDOH cofactors. Medical researchers are conditioned to see and accept ‘race’ as explaining difference.

However, Black patients in the general population endured **(1)** deprivations that aggravate tissue injury, **(2)** “race corrected” eGFRs that delayed CKD diagnosis, and **(3)** systematically delayed CKD care. **HOWIE** also showed that FSGS histology can evolve [461], so worse histology in Black patients is hardly surprising, given the extreme exposures (see **Appendix: 5.6 Case study: North Carolina**). Merging the arbitrary MCD and FSGS categories (the two data columns at far left) into a single data column (far right), **Fig 21**, suggests similar <u>overall</u> FSGS frequency for all patients.

#### 5.5.2 APOL1 Geography

##### APOL1 in the US

Racialization of APOL1 follows virtual absence of *APOL1* RVs outside the West African population. Because “…most current knowledge about the role of *APOL1* genetic variants on diseases are extrapolations from studies on… mainly African Americans in the USA” [462], APOL1 research is overwhelmed and confounded by US ‘racial’ illusions and systemic racism. Our research directly addresses APOL1 in the US. Comparing APOL1 outcomes in other regions offers insights.

##### APOL1 in Africa

In 2014, an “update” on *APOL1* kidney risk alleles predicted the rate of CKD would be “…much higher in sub-Saharan Africa, where the G1 and G2 alleles are found in most of the populations surveyed, rates of HIV infection are among the highest in the world, and treatment options are limited” [463].

A South-African study of HIV-associated nephropathy (HIVAN) in black patients concluded “…antiretroviral therapy-naïve South-African blacks with two *APOL1* risk alleles are at very high risk for developing HIV-associated nephropathy” [464]. However, antiretroviral therapy (ART) markedly reduces HIVAN, and the study also noted that “All HIV-positive individuals with CKD were ART naïve and 75% of the HIV-positive control group was receiving ART”.

In Nigeria and Ghana, adjusted odds ratio for CKD in patients with two copies of the RV alleles (i.e., *APOL1* HR genotype) versus zero or one copy was only 1.25 (95% confidence interval [CI], 1.11 to 1.40) [465], well <u>below</u> the US ratio of three times greater CKD in Black than White patients [465]. That study also noted, “The odds ratio of approximately 2 for focal segmental glomerulosclerosis among participants with two APOL1 risk alleles in this study is modest in comparison with the previously reported odds ratio of approximately 10 among Black Americans. The reason for this difference is unclear”. Studies overlook the medicontext of US structural racism as, perhaps, more salient than risk factors prevalent in Ghana and Nigeria.

In Ghana, Nigeria, and South Africa after English colonial rule, South African apartheid kept the most severe racialization. An *APOL1* RV pigmentary gradient may proxy for worst access to healthcare under South African colorism [466], as for structural racism in the US, with *APOL1* associations “harmonized” to international ‘racial’ categories (see **Section 3.3.3(c) Pharmaceuticals “harmonizing” ‘race’**).

##### APOL1 in Europe

A European study of *APOL1* variants in kidney transplantation patients found no instances of the RVs reported in African Americans and only one haplotype occurring in more than 1% overall <u>and</u> more often in the CKD category G5 cohort [5]. The authors concluded that APOL1 does not likely contribute much to CKD risk, as the difference was attributed to a single base-pair deletion with a homozygous condition. Further, the authors performed 16 hypothesis tests in that table alone, creating a 56% chance of at least one significant result due to chance alone. This suggests that their evidence indicating an effect of APOL1 outside of West African ancestral groups is scant, at best, and may be more plausibly explained as an artifact of repeat hypothesis testing.

##### APOL1 in Taiwan

Taiwan has one of the highest rates of kidney disease. A Taiwanese study of rare missense variants for CKD and ESKD susceptibility found none of the *APOL1* RVs [6].

### 5.6 Case study: North Carolina

#### Environmental racism

“Environmental racism” is one key to understanding kidney health [467]. The term first appeared in the medical literature in 1991 [468], and in only seven articles through 2017. From 2018, more than 80 PubMed-indexed articles used the term to report toxic exposures rooted in structural racism and persistent ‘racial’ segregation. North Carolina (NC) was ground zero in the fight against environmental racism. Sources of water and air pollution were often built in neighborhoods of poor and Black people, including toxic landfills [469,470], “concentrated animal feeding operations (CAFOs)” [471], and the coal-burning power plant of the University of North Carolina at Chapel Hill [472].

> In NC and elsewhere, racial minorities and low-income individuals are found to be underserved with respect to municipal water and sewer services. This is partially a result of institutional racism ingrained in municipal zoning practices known as “underbounding,” in which the water and sewer service boundaries have historically been intentionally underdrawn in the U.S. South to exclude minorities. As a result, these communities are forced to rely on private wells, despite the proximity of a municipal water system; this places them at a higher risk for chemical exposure via water, with the corresponding health risks. [473]

“NC has one of the highest proportions of the population relying on federally unregulated private well water…” [474], and “…areas in the “low % black” category have the highest odds of water service: odds of water service in these areas are 85% higher than in census blocks that are 100% black (p<0.001)” [475]. Heavy metals (e.g., lead, inorganic arsenic, and manganese) are common contaminants [474,476], and there is ‘racial’ disparity in whether a well is even tested [477].

However, the problem is nationwide: “…households headed by people of color are almost 35% more likely to lack piped water as compared to white, non-Hispanic households” [478]. “[C]hildren in homes relying on private wells have 25% increased odds (95% CI 6.2 to 48%, *P* < 0.01) of elevated blood Pb, compared with children in houses served by a community water system that is regulated under the Safe Drinking Water Act” [479].

#### Convenient associations hide inconvenient facts

North Carolinian authors defined CG as a ‘race’-based genetic disorder because, they said, “No patients had known toxin exposure” [89]. Others enthused about the *MYH9* gene for KF in African Americans [90] but moved on, for now, to *APOL1* [271].

**Ethicist Roberts** noted:

> The concept of biological race is useful to explain away inequities that are caused by differences of political power…. The persistent belief that these blatant inequalities of wealth, of incarceration, of income, of health, of education, in the United States, stem from innate differences—it’s so much easier to believe that than to actually face the continuing impact of racism in our society. [280]

Despite our research, medical journals may still promote eGFR, FSGS, and APOL1 to Black Americans and others, which is disappointing but not surprising. Historical ethicists can judge whether late-20th-century conclusions of CG and *MYH9* met ethical standards of that time, whether medical journal policies blinded researchers to their historical context, and whether it was just plain BS (bad science) of biological ‘race’ [229]. A more important observation is that flawed “peer” review allowed distinguished colleagues far from NC to amplify that BS.

Biological ‘race’ is an illusion, and “peer” reviewed silence is consent. Many of today’s North Carolinian researchers center ‘racial’ disparities in lingering ‘racial’ segregation and structural racism. The benefits for children may not be known for decades, but remedying an injustice started a natural experiment: “Bordering the wealthy town of Apex, North Carolina, is a majority African-American neighborhood… without town water service, relying on private wells” [480], but “A decision by Apex to annex the area and extend water service…” [481] might reduce incidence of pre-natal and post-natal kidney injury and, decades from now, lead to reduced late development of CKD and KF.

As we noted (in **Section 3.3.2 Racism’s genome-wide associations**), genes in linkage disequilibrium with pigmentary genes are more likely proxies for the harms from racism that better explain the data patterns. Bioethical oversight of ‘race’ could require broader hypotheses and data collection to get to the roots of ‘racial’ disparities.

### 5.7 Silence on ‘race’

**Alana Lentin, Professor of Cultural and Social Analysis at Western Sydney University**, described the roots of European silence on ‘race’ after the Holocaust [297]. By forbidding discussion of ‘race’ and substituting euphemisms like “ethnocentrism”, Europe supports our well-meaning colleagues in US medical publishing who have long enforced similar silence on ‘race’. Medical editors and journals presented a double standard in reluctance to remove ‘race’ from medical equations: ‘race’ doesn’t count when it’s about discussing ‘racial’ disparities and the history of racism, but to provide medical care, it is essential and unignorable. Agreeing to disagree, geneticists and anthropologists took two sides of the same coin—racism and ethnocentrism, advancing the segue from unacceptable racism to “acceptable” ethnocentrism. Three excerpts from **Lentin**:

**(a) “…a central ordering principle…”**

> The race critical perspective… counters the predominantly held idea that any talk of race should be silenced because it has been proven to be a scientifically unviable concept that in the wrong hands leads to evil. Accepting that race is socially constructed…, it is nevertheless understood that race has become a central ordering principle of modern western societies. This is because race has produced rac*ism*, which today… manifests itself in a variety of institutional and societal discriminations, stereotypings, and injustices that in many instances… are dissociated from their origins in the race projects of the eighteenth and nineteenth centuries (slavery, colonialism, eugenics, genocide …). [297]

**(b) “…ethnocentrism to be used in the place of racism.”**

> Race… was literally banished from publicly acceptable discourse, particularly in continental western Europe. For Goldberg (2006: 336), after the *Shoah*, ‘race is to have no social place, no explicit markings. It is to be excised from any characterising of human conditions, relations, formations.’ This does not mean, however, that it ceases to be implicit. Just as anthropologists and geneticists agreed to disagree, leaving the latter free to develop theories of racially underpinned intelligence and proneness to disease, race was not uttered but could not be unthought….
>
> Anthropologists… argued for terms such as ethnicity and culture to replace that of race, and for ethnocentrism to be used in the place of racism. It was recognized that the horrors of the *Shoah*, steeped in the taint of race, were not dissociable from the difference made evident by the appearance of non-whites in the European midst. Yet it was this very association with the *Shoah* that led to the products of colonial rule (the postcolonial immigrants) becoming ‘ethnic minorities’ and ‘cultural groups’, rather than races.
>
> The evident outcome of the replacement of race with ‘culture’, ‘ethnicity’ or, even more euphemistically, ‘background’ is the consequent banishing of *racism*. It is of course correct that race should not be reinstated to its pre-war status. Yet, … race is given meaning by the persistence of racism. [297]

**(c) “…transformed race into ‘identity’.”**

> Unable/unwilling to speak about the effects of race, Europe… has transformed race into ‘identity’. In other words, race is no longer something that is imposed upon subjugated others. By anaesthetizing race and labelling it ‘ethnicity’ or ‘culture’, it becomes something that is possessed, rather than something that is unwillingly acquired… The badge of race is not worn… out of choice. Even where race is proudly reappropriated, it is as a result of racism that the need to organize around a far from authentic racial identity is felt. In the contemporary context, the racialization of religious affiliation, most notably that of Muslims, is a clear example of how an oppositional shared identity can congeal and harden in reaction to the presumed immutable otherness of a shared faith….
>
> In other words, it is difference per se that is problematized by the various debates and policies that frame what is no longer known as race in Europe. The spectre of race lingers in the sense of discomfort that impels us to find solutions to the ‘living together’ (Touraine, 2000) of culturally (racially) different, and it is implied, incompatible groups. Moreover, this is portrayed by the liberal establishment as something that is called for and expected by monolithically named ‘minority’ groups. The framing of the grievances of the racialized as identity politics (Lentin, 2004, 2005), means that what have mainly been unsuccessful policies are put down to the recognition claims (Taylor, 1994) of these marginalized ‘minorities’. The call of the racialized to stand against racism is translated into a call to recognize (my) identity. Of course, this entirely overlooks why identity or race as a Du Boisian badge takes on the importance it does in the first place….
>
> …by refusing to see race, by covering it over with alternative signifiers and therefore, most crucially, blinding itself to its consequences, Europe sends a message: race is not our problem, it is not ours. It is yours. [297]

During a discussion of ‘modern race’ and the need for openness, **Medical ethicist Charmaine Royal** noted,

> Racism created race and is the root cause of the associated inequities and disparities we see today. This narrative must replace inaccurate narratives about the relationship between race and racism and the reasons for differences in health and other outcomes among racialized groups….
>
> …more than 50% of the U.S. population believes that there are biological human races. Furthermore, research has shown that biological race thinking influences how we do scientific research, how we provide health care, and how we live in society. To achieve health equity, we must dismantle beliefs in the existence of biological human races and racial hierarchies that underlie and perpetuate structural inequalities and health disparities. [482]

### 5.8 Black and “NOT Black”

Black Americans are especially aware of inconsistent ‘racial’ categories, for example when traveling abroad and under immigrant bosses at work and in training, **Fig 22**.

**Fig 22.**
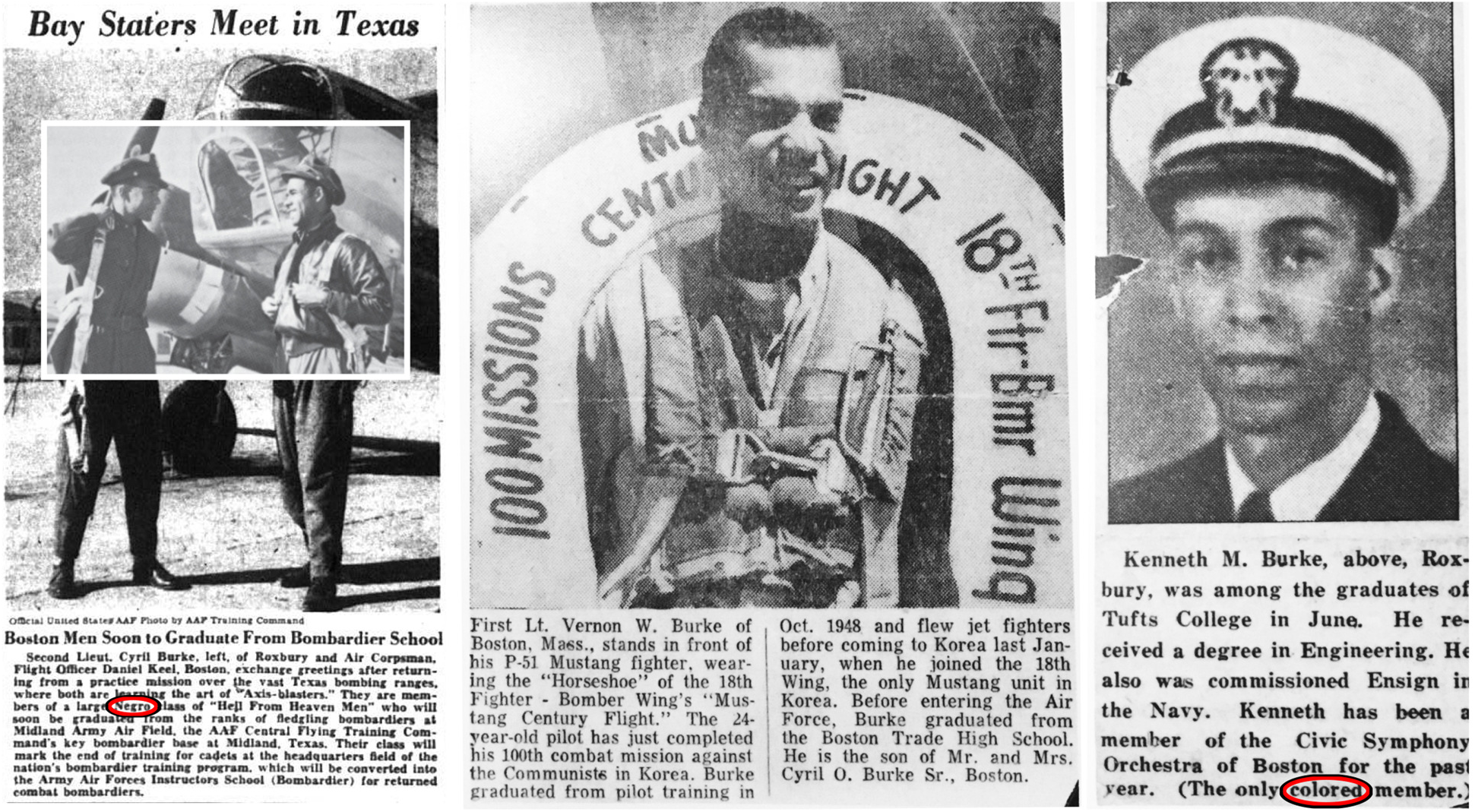
Irresistible inconsistency meets immovable ‘racial’ beliefs—. Consider three brothers, family of one of the authors (cob3). During segregation, newspapers reported “Negro” and “colored”. In 1960s [222], self-identification embraced “Black” and “African American”. In 1990s, senior academic physicians from Canada and South Africa <u>insisted</u>, respectively, on “White” and “NOT black… There is no more racism and no more antisemitism, either!” A US-born physician <u>insisted</u> on thiazides as best treatment, but the patient (cob3) rejected ‘race’-based care for standard “renoprotective” therapy. Various ethicists could insist that biological ‘race’ is an illusion and hold academic medical research to modern ethical standards against misuse of ‘race’.

## ACKNOWLEDGMENTS

In memory of Rear Admiral W. Norman Johnson and many others who endured kidney failure after being prescribed nephrotoxic drugs that might have been avoided with early warning and caution, and of the Rev. Dr. Canon Cyril C. Burke, Sr., who taught ethics and whose final medical care was complicated by ‘race’.

The authors gratefully acknowledge Edward Feller, MD, FACP, FACG, Ruth Levy Guyer, PhD, James M. Gilchrist, Professor Emeritus of Neurology at Southern Illinois University School of Medicine, and anonymous colleagues for critical review of a draft of this article, and Joseph J. Fins, MD, MACP, FRCP, and John C. Kotelly, PhD, US Air Force mathematician (ret.), for their insights.

## STATEMENTS AND DECLARATIONS

### Declaration of conflicting interest

The authors declared no potential conflicts of interest with respect to the research, authorship, and/or publication of this article.

### Funding statement

The authors received no funding for this work.

### Data availability

All relevant data are provided in the primary sources.

### ABBREVIATIONS

AKI: acute kidney injury
APOL1: apolipoprotein L1
BS: bad science
CG: collapsing glomerulopathy
CKD: chronic kidney disease
COI: conflict of interest
COVID: coronavirus disease
eGFR: estimated GFR
EU: European Union
FSGS: focal segmental glomerulosclerosis
FDA: Food and Drug Administration
GFR: glomerular filtration rate
GWAS: genome-wide association study
HR: high risk
IRB: institutional review board
LR: low risk
KF: kidney failure
MDRD: Modification of Diet in Renal Disease
NASEM: National Academies of Sciences Engineering and Medicine
NC: North Carolina
OR: odds ratio
PCT: pragmatic clinical trials
PRS: polygenic risk score
RR: relative risk
RV: risk variant
SDOH: social determinants of health
sIRB: single IRB
srPRS: structural racism polygenic risk score
UK: United Kingdom
US: United States,
VA: Veterans Administration

## Notes

### Competing Interest Statement

The authors have declared no competing interest.

### Author Declarations

The study used ONLY openly available human data that were originally published in the three source articles.

### Summary of Updates

(1) Researched and rewrote Section 3.2.6 Focal segmental glomerulosclerosis, (2) New Figs 0, 16, 19, 21 (2) Added Appendices: 5.5 Medicontextual FSGS and APOL1, 5.6 Case, study: North Carolina, and 5.7 Silence on 'Race', (3) Improved formatting of block quotations, (4) Revised Figs 18, 20a, 20b, (5) Minor revisions throughout the text.

